# rendehR: a cohort-first, provenance-aware, agentic-ready pipeline for harmonizing multi- source electronic health records

**DOI:** 10.64898/2026.11.19.26367722

**Authors:** Marzieh Mussavi Rizi, Vladyslav Shein, Abel Torres-Espin

**Author notes:** Corresponding author: Abel Torres-Espin, PhD School of Public Health Sciences University of Waterloo, Waterloo, Ontario, Canada.

## Abstract

**Objective:** Public electronic health record (EHR) databases differ in schema, identifiers, coding systems, units, and event structure, making disease-specific studies across multiple sources difficult to construct and audit. We developed rendehR, a computational pipeline and R package for cohort-centered extraction and harmonization of multi-source EHR data.

**Materials and Methods:** rendehR treats cohort definition as an explicit executable component of dataset construction. Cohorts can be defined using diagnosis codes, identifiers, R callbacks, or source-specific SQL. After cohort selection, linked source tables are restricted to the selected population and can be retained as cohort-specific Parquet datasets or loaded into a persistent database. Clinical concepts are defined using source-aware mappings, generated, or curated SQL and are harmonized into flat, time-series, and duration representations. Configuration, executable SQL, cohort metadata, concept-origin metadata, and source-level values are retained to support inspection and traceability. In the current version rendehR provides interactive R assistant wizards for workflow configuration, a Shiny application for patient-level data visualization, three agent skills for AI-assisted workflow execution and extension, and an MCP server exposing nine read- only tools for AI-assisted configuration, concept discovery, and cohort exploration. The current implementation supports MIMIC-III, MIMIC-IV, and eICU-CRD. We demonstrate rendehR on a reference cohort of spinal cord injury and disease (SCID).

**Results:** Using the reference cohort, rendehR identified 4,320, 39,850, and 2,343 qualifying hospital admissions in MIMIC-III, MIMIC-IV, and eICU-CRD, respectively. Patient-level extraction expanded these cohorts to 5,725, 104,988, and 2,823 hospital admissions, with 5,983, 17,405, and 3,441 associated ICU stays, respectively. Applying the more complex Sepsis-3 case identification, rendehR identified and extracted 34,460, 41,296, and 47,035 qualifying ICU stays across the same databases, respectively. The current concept library includes 22 flat, 123 time- series, and 75 duration harmonized concepts.

**Conclusion:** rendehR provides a cohort-centered framework for constructing disease-specific datasets from multiple heterogeneous EHR sources while preserving the source-specific logic used to define cohorts and clinical concepts. The package supports reusable cohort artifacts, inspectable SQL-based concept construction, harmonized analytical representations, and traceable study- specific data pipelines.

**Graphical Abstract:** A cohort-first, provenance-aware, agentic-ready pipeline for harmonizing multi-source EHR data

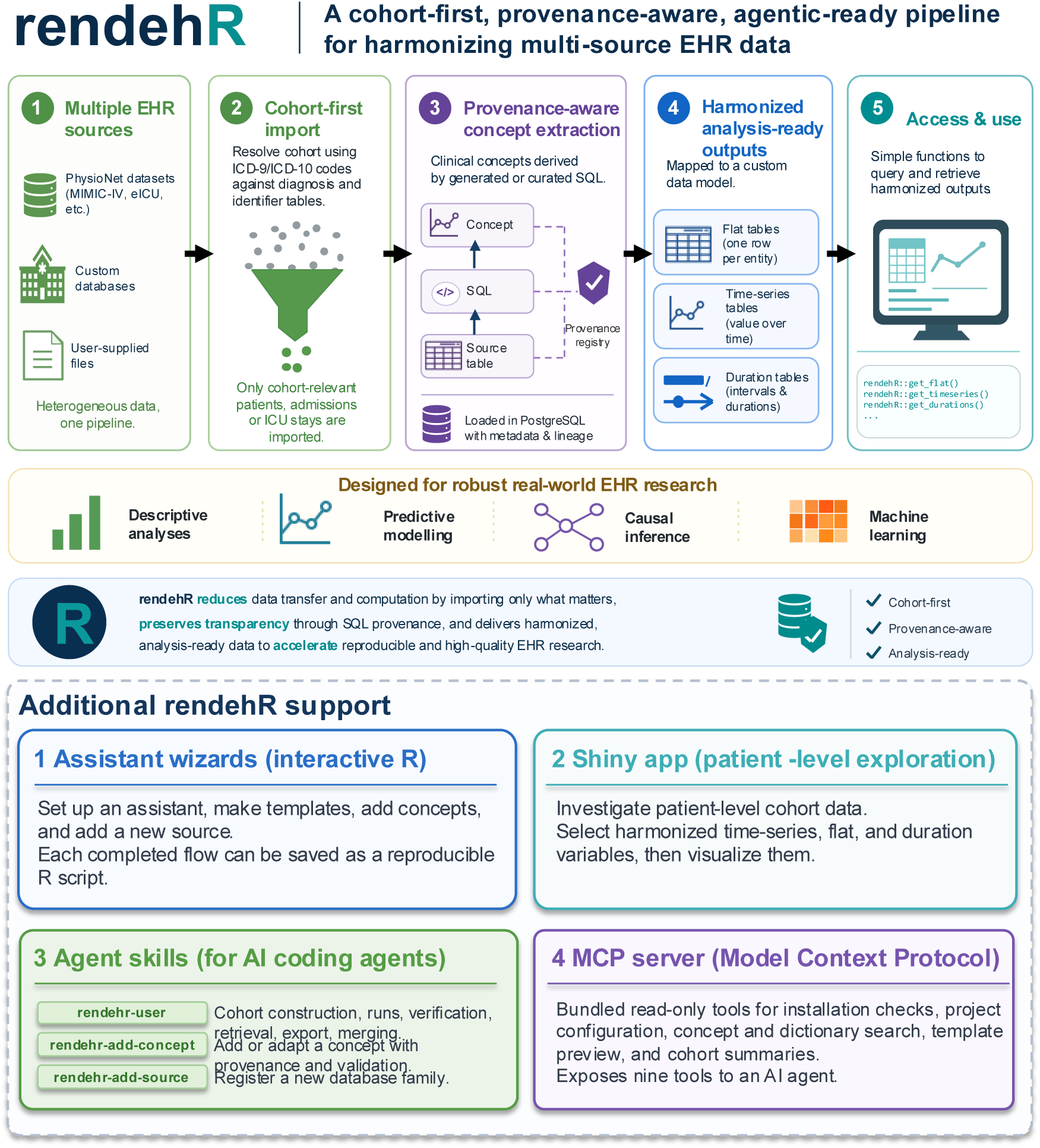

## 1. Introduction

Electronic health records (EHRs) are an important resource for clinical research. Public resources such as the Medical Information Mart for Intensive Care III (MIMIC-III) [1], MIMIC-IV [2], and the eICU Collaborative Research Database [3] provide detailed de-identified clinical data and have become widely used in observational studies and machine learning benchmarks [4–6]. They contain demographics, diagnoses, procedures, medications, laboratory measurements, and physiological observations, but represent this information differently across sources.

EHRs provide an opportunity for multi-source study; however, such studies require source-specific decisions before patients and clinical variables can be represented in a common analysis dataset. Medical concept extraction, for instance, involves technical and clinical choices: which source tables and item identifiers to use, how to reconcile units and timestamps, and which plausibility rules to apply. These choices shape how a concept appears in the final dataset and affect how downstream analyses can be interpreted. Harmonizing medical concepts, therefore requires balancing between enabling cross-source analysis and preserving clinically meaningful source- specific details [7].

A range of software tools in Python and R have been developed to facilitate the extraction, processing and harmonization of EHR data. MIMIC-Extract [8] and FIDDLE [9] provide preprocessing pipelines for structured EHR data, focused on features construction for downstream prediction tasks. The MIMIC-Code and eICU-Code repositories provide reusable SQL implementations of derived clinical variables [10,11]. In R, rEHR [12] and rcprd [13] provide database-backed workflows for extraction and processing of large primary-care EHR data, while dxpr [14] provides tools for diagnosis and procedure code processing. Ricu provides a unified interface to multiple ICU databases, including MIMIC and eICU-CRD [15] by separating source- agnostic clinical concepts from the source-specific mappings required to extract them. The Python BlendedICU library harmonizes multiple ICU databases into a common dataset and maps the harmonized representation to the Observational Medical Outcomes Partnership (OMOP) [16] common data format [17].

Existing tools generally harmonize the full source database first and extract the study population afterward. However, a disease-specific study typically starts with identifying the population defined by the research question and the clinical variables extracted afterward. Cohort identification and variable construction may require source-specific diagnosis mappings or a more complex executable phenotype. A disease-specific workflow therefore needs to support cohort extraction while preserving the source-specific definitions and transformations that produced the analytical dataset.

We developed rendehR to support a workflow that integrates with the R ecosystem, scales across data sources and harmonized concepts, and is designed for cohort-first, provenance-aware applications. rendehR treats cohort definition as an explicit executable component of dataset construction. Cohorts can be specified using diagnosis codes, identifiers, or source-specific SQL, allowing definitions that range from code-based phenotypes to algorithms such as sepsis. After cohort membership is determined, source data is restricted to the selected population, and clinical concepts are derived using source-specific SQL. Simple concepts can be generated from configuration files, while more complex definitions can use curated SQL, including adaptations of community implementations where appropriate. The package retains the SQL and provenance associated with cohort and concept construction, and links these definitions to persistent, derived tables and harmonized outputs.

Given the growing use of large language models and agentic approaches in clinical data harmonization, including clinical concept mapping, automated SQL generation, and multi-agent data transformation workflows [18–20], we extended rendehR to support AI-assisted workflows. We added three agent skills for cohort construction, concept development, and integration of new data sources, along with an MCP server exposing nine read-only tools for accessing project configurations, concept definitions, source dictionaries, and cohort summaries.

We evaluate rendehR using MIMIC-III, MIMIC-IV, and eICU-CRD. Spinal cord injury is used as a case example to demonstrate use and evaluate cohort construction across different diagnosis and identifier structures, whereas sepsis is used to evaluate SQL-defined cohort selection based on a multivariable clinical phenotype. We also compare selected clinical concepts with established reference implementations, quantify the data retained after cohort selection, and demonstrate construction of an analysis dataset from the harmonized outputs.

## 2. Materials and Methods

### 2.1 Data sources and software environment

rendehR currently supports three public EHR databases available through PhysioNet: MIMIC-III version 1.4 [21], MIMIC-IV version 3.1 [2,22], and eICU-CRD version 2.0 [23]. MIMIC-III contains 58,976 hospital admissions and 53,423 ICU stays from Beth Israel Deaconess Medical Center between 2001 and 2012. MIMIC-IV expands this resource, containing 546,028 hospital admissions and 94,458 ICU stays from 223,452 unique individuals. eICU-CRD is a multi-center source, with more than 139,000 patients and 200,000 ICU stays from 208 hospitals across the United States. These databases contain de-identified demographics, diagnoses, medications, laboratory measurements, physiological observations, procedures, and clinical outcomes, but differ in their relational structure, identifier hierarchies, diagnosis representations, clinical item dictionaries, and organization of hospital- and ICU-level events.

Access to the full databases is provided through PhysioNet platform [24] and requires completion of the corresponding credentialing and data-use requirements. Once users obtain the required credentials and access permissions from PhysioNet, they can use rendehR to download and process the approved EHR datasets. The package also provides preconfigured workflow for MIMIC-III Demo version 1.4 [25], MIMIC-IV Demo version 2.2 [26], and eICU Demo version 2.0.1[27]. The demo datasets are publicly available and can be used without credentials.

rendehR is implemented in R (>=4.1) [28]. DuckDB [29] is used to query raw source files, execute cohort-selection logic, resolve cohort identifiers, and filter source tables before construction of the cohort-specific dataset. Cohort-restricted datasets are written to Apache Parquet [30] and subsequently loaded into the database backend. PostgreSQL [31] is currently used for the full downstream workflow, including curated concept execution, harmonization, multi-source merging, and the primary data-access interfaces. DuckDB can additionally persist cohort databases and execute supported concepts; full DuckDB backend support is planned for version 2.

### 2.2 rendehR workflow

rendehR organizes EHR dataset construction around the study cohort (Figure 1). Source configuration defines the raw tables, data types, variable mappings, and patient, hospital-admission, and ICU-stay identifiers for each database. The cohort definition is executed first; source tables are then restricted to the selected patients. Cohort-restricted data can then be stored as Parquet files or loaded into a persistent PostgreSQL database.

**Figure 1.**
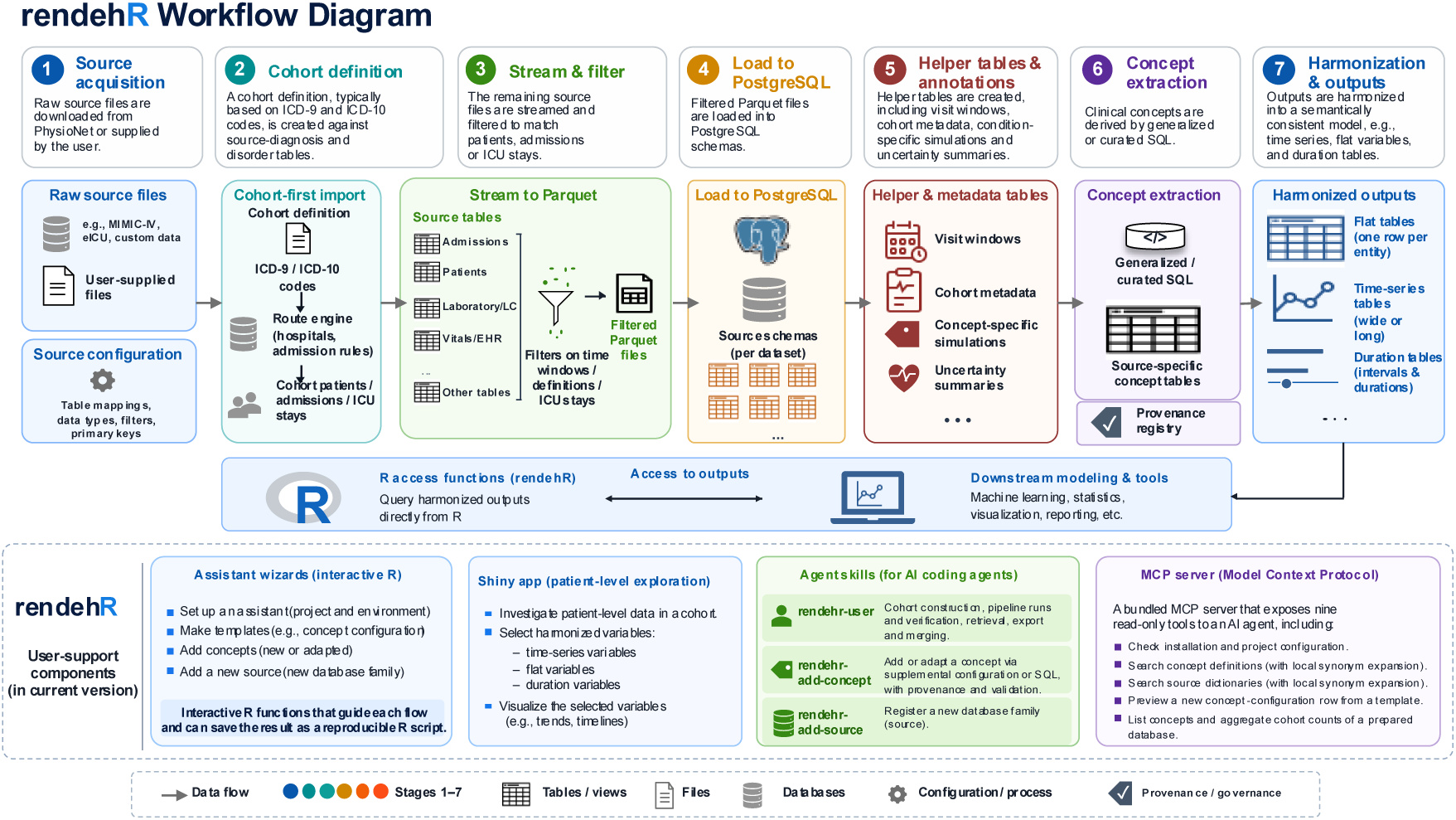
rendehR workflow. (1) Source acquisition; (2–3) cohort definition and extraction; (4-5) loading of cohort-restricted source data into the database backend and creation of helper tables; (6) construction of source-specific clinical concepts; (7) harmonization into flat, time-series, and duration representations; and merging of source-specific outputs for downstream access and analysis. Interactive assistant wizards support workflow configuration and R script generation, while a Shiny app enables patient-level visualization of harmonized data.

Next, clinical concepts are constructed from source-specific definitions. rendehR classifies concepts into two general categories, atomic and complex, based on the complexity of their extraction requirements. Extracted concept tables are then harmonized into flat, time-series, and duration representations. Each stage produces persistent intermediate outputs that can be inspected independently and rerun without reconstructing the entire pipeline.

### 2.3 Cohort definitions and extraction

rendehR supports several cohort-definition methods. A cohort can be specified using diagnosis- code resources, a set of patients, hospital-admission, or ICU-stay identifiers, an R callback, or user-supplied SQL. SQL cohort definitions are executed in DuckDB against typed views of the raw source files. More complex phenotypes can use an ordered sequence of setup SQL statements before the final cohort query runs.

The current version of rendehR includes SCID and Sepsis-3 as packaged reference cohorts. They illustrate two different cohort-definition mechanisms: diagnosis-based selection and an executable clinical phenotype defined in SQL. For diagnosis-based selection, users provide a dictionary of ICD-9-CM and ICD-10-CM codes, which can be supplied as CSV or RDS files. The dictionary must contain the diagnosis codes and their corresponding ICD versions, and codes can be provided with or without decimal points. The SCID reference dictionary additionally includes long descriptions and related diagnostic categories. For SQL-based cohort selection, users provide the path to a SQL file or a series of SQL files containing cohort-selection queries written in DuckDB- compatible SQL. rendehR then executes these queries against the source data to identify the qualifying identifiers of the cohort, as demonstrated by the Sepsis-3 phenotype.

The output of cohort selection is then mapped to the identifier hierarchy defined for each source. The identifier returned by the cohort definition determines the selection grain, whereas a separate extraction-scope argument determines whether broader linked context is retained (Figure 2). For example, the granularity of sepsis cohort is at ICU-stay level because the cohort is identifies qualifying ICU stays based on Sepsis-3 criteria [32]. The user can keep only those stays or expand the extraction to the corresponding hospital admission (and include non-septic ICU stays) or full patient record available for a patient once was diagnosed with sepsis. A diagnosis-code-based cohort such as spinal cord injury and disease (SCID), on the other hand, is commonly identified at the hospital-admission level. In that case, records from ICU stays linked to the selected admission are kept as part of that admission-level context, while extraction can be expanded further to include the patient’s broader record when required.

**Figure 2.**
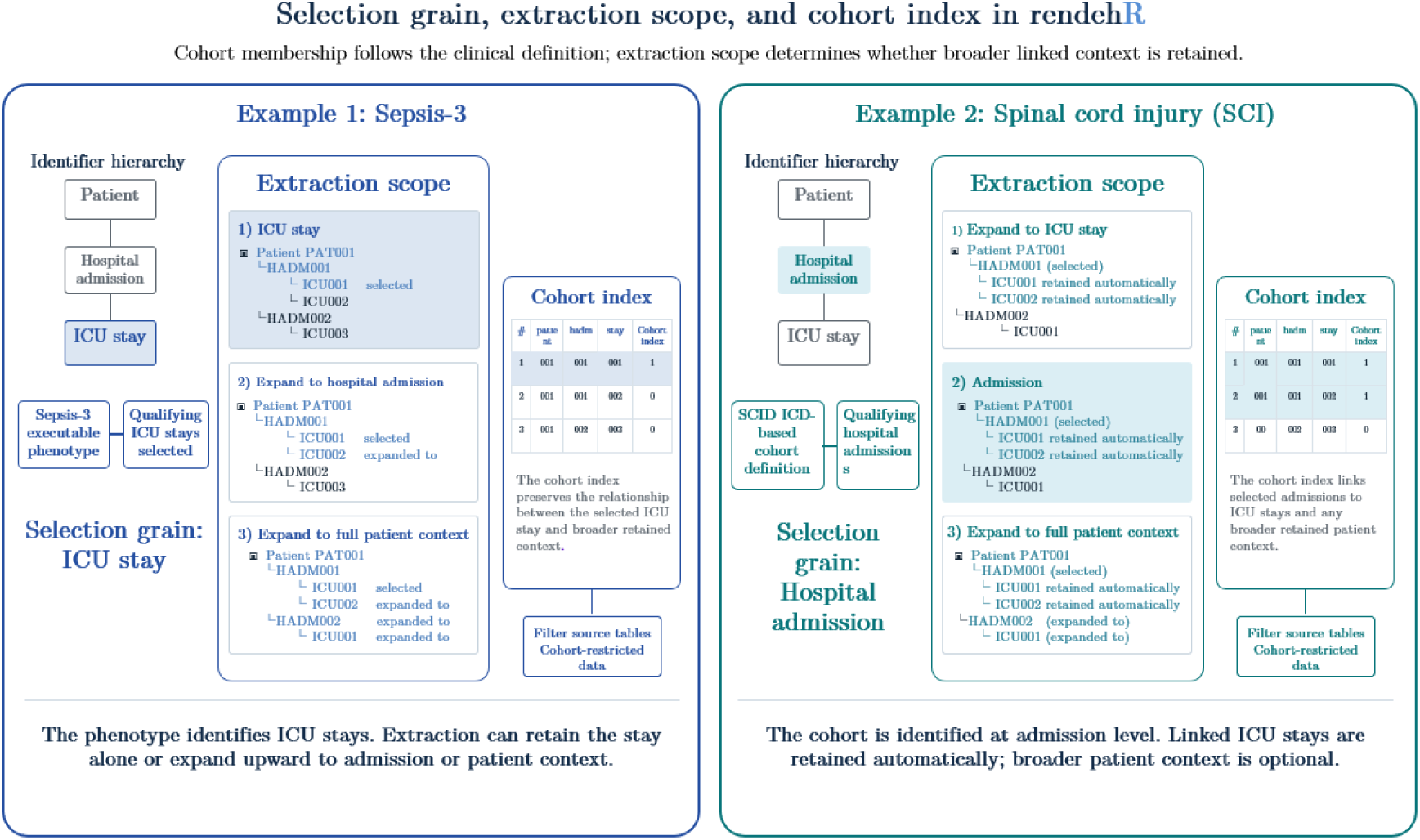
Cohort selection grain, extraction scope, and cohort-index. The cohort definition selects the granularity at which cohort eligibility is established, while the extraction scope determines the broader linked context retained. The cohort index records patient-, admission-, and ICU-level identifiers included after scope expansion. SCID and Sepsis-3 illustrate admission-level and ICU- stay-level selection, respectively.

When the extraction scope is expanded, the linked patient-, admission-, and ICU-level identifiers are recorded in a cohort index. This index identifies which records belong to the selected cohort context and is used to filter downstream source tables during cohort-specific dataset construction.

Once cohort membership and extraction scope have been resolved, clinical tables are filtered to the selected cohort before downstream concept construction. Reference tables that are not patient- specific are kept in their original form. The cohort-restricted tables are converted to Parquet files which the user can choose to keep. This Parquet output is itself a reusable research artifact. It provides a portable, cohort-specific representation of the source EHR that can be used independently or as input to the downstream rendehR concept and harmonization workflow.

### 2.4 Extraction of clinical concepts

Once the cohort database is built, clinical variables are defined through source-aware concept specifications. rendehR follows the source-agnostic concept principle used by ricu [15] by separating the clinical meaning of a variable from the source-specific instructions needed to extract it. A source-specific recipe specifies the source table, item identifiers or matching expressions, timestamp and value fields, units, transformations, and optional value-range rules. rendehR differs from ricu in how these definitions are executed and retained. In rendehR, source-specific concept definitions are compiled into inspectable SQL, executed against the cohort-specific database, and retained together with the resulting concept tables and provenance information.

rendehR categorizes concepts into two general categories, atomic and complex, based on how they are extracted. Atomic concepts are variables that can be extracted from a source table using source mapping and simple transformation rules, without requiring a multi-stage clinical derivation. For atomic time-series and duration concepts, these specifications are stored in configuration files and translated into SQL before execution. The generated SQL is written to source-specific files, so the implemented definition can be inspected directly. More complex concepts, those requiring joins across multiple tables, temporal logic, or multi-stage derivation, are implemented using curated SQL. Users can override existing concepts by editing the original configuration file or supplying a new one from the provided template. Additional atomic concepts can be added through configuration files; complex concepts can be added with user SQL.

### 2.5 Provenance and traceability

To make each concept and cohort traceable, rendehR records provenance at each stage of cohort and concept construction. Cohort metadata captures the selection route, identifier level, extraction scope, cohort counts, cohort identifiers, source release, and build time. Table-level reports are also generated during cohort construction. The cohort hierarchy links patient-, hospital-admission-, and ICU-stay identifiers, defining the extraction context. Cohorts trace back from the packaged ICD definition or SQL definitions, through source-specific qualifying identifiers and the cohort hierarchy, to the metadata and table reports.

Concept provenance is stored in four artifacts; a) Concept configuration files record source mappings, units, value ranges, and extraction item identifiers or matching rules; b) SQL files contain the executable definition, whether generated or curated; c) Concept tables hold the raw data alongside source values and identifiers for each source; d) The registry stores attribution and build metadata, with each entry listing concept name, output schema, origin, reference or URL, and build time. A concept can be traced by starting from its harmonized name, finding the source mapping or SQL, then locating the source-specific table and its registry entry.

### 2.6 Harmonized analytical representations

After source-specific concepts are constructed, rendehR harmonizes them into three output types: flat, time-series, and duration tables. Flat tables contain variables which are constant at the relevant analysis level, such as demographics, comorbidities, outcomes, or summarized clinical characteristics. Time-series tables contain repeated measurements in long format, with the harmonized concept name, value, unit, source identifiers, and measurement time. For events with defined start and end times, duration tables represent interval-valued events, such as treatments or medical ingredient exposures.

Time-series and duration observations are aligned relative to hospital admission using elapsed time in minutes. When ICU timing is available, ICU-relative timing and ICU length-of-stay information are also retained. The same concept tables, therefore, support hospital-relative or ICU-relative analyses without redefining the underlying concept.

Harmonized tables can then be combined across supported sources while retaining the source database and native identifiers. Access functions provide long-format data directly or construct wide representations using user-specified time bins and aggregation functions. These outputs are intended to reduce source-specific extraction work. At the current version, tasks such as missing- data handling and feature construction stay with the investigator.

### 2.7 Verification and validation

rendehR implement both technical verification and concept creation validation. Technical verification checks whether the pipeline produces the expected files, schemas, and tables at each major stage, lowering the risk that a pipeline failure slips silently into the analysis datasets. Five functions cover the main stages: the download process, database generation, and cohort extraction and harmonization.

Concept-level validation compares selected concepts against an established reference, the ricu package. This comparison shows where rendehR and the reference agree, and whether any differences are expected given the study definition. Using the spinal cord injury cohort across all three sources, we compared source-specific mappings, units, timestamps, row counts, and value distributions for each selected concept.

We compared value distributions for all time-series concepts shared by rendehR and ricu. For each concept and data source, we compared values from the same hospital admissions using the two- sample Kolmogorov–Smirnov statistic D [33,34]. This statistic measures the largest vertical distance between the two empirical cumulative distribution functions, ranging from 0 for identical distributions to 1 for complete separation. We first excluded rendehR values outside ricu’s plausibility limits to match ricu’s built-in filtering, then calculated D using all values for each concept.

Comparisons involved up to one million values, so even very small differences could be statistically significant. We therefore used D as a descriptive effect size and predefined concordance as D ≤ 0.05. Asymptotic p-values are reported for completeness. Concepts for which the packages intentionally use different source items were documented in advance, together with the reasons, and are summarized separately. For each source, we report the number of concepts, the mean and range of D, and the concept with the largest distance. Results for individual concepts, including p-values, are provided in Supplementary Table 6 and 7.

Validation covered sex, age, length of stay, and in-hospital mortality as flat variables, along with all 65 common time series variables and five exposure variables as examples of duration variables. Clinical plausibility summaries report observed units, value ranges, selected quantiles, missingness, observation counts, unique subjects or visits, and temporal distributions where applicable. Where the concept configuration specifies expected units or value ranges, observed values are checked against them. These summaries are then reviewed to flag unexpected values, units, or distributions. Agreement in mapped items, matched values, and summary measures of numerical differences is reported in Section 3.4.

### 2-8 Interactive tools and AI agent support

Three components in the current version of rendehR supports users. Assistant wizards are interactive R functions for setting up an assistant, making templates, adding concepts, and adding a new source. Once a flow is complete, the result can be saved as an R script. Moreover, to investigate patient-level data in a cohort, a complementary rendehR Shiny app is provided in the package, allowing users to select the harmonized time-series, flat, and duration variables that exist for a patient and visualize them.

The package also ships three agent skills: rendehR-user (cohort construction, pipeline runs and verification, retrieval, export and merging), rendehR-add-concept (adding or adapting a concept through supplemental configuration or SQL, with provenance and validation steps) and rendehR- add-source (registering a new database family). Skills are instruction sets that an AI coding agent loads on demand; depending on the request, the agent returns reproducible R code or runs it for the user.

The third component is a Model Context Protocol (MCP) server bundled with the package, which exposes nine read-only tools to an AI agent: checking the installation and project configuration, searching the packaged concept definitions and the source dictionaries (with local synonym expansion), previewing a new concept-configuration row from an existing template, and listing the concepts and aggregate cohort counts of a prepared database.

## 3. Results

### 3.1 Case examples of cohort construction across sources

As use-case examples, we used rendehR to construct a spinal cord injury and disease (SCID) and sepsis cohorts across three supported EHR sources (MIMIC-III, MIMIC-IV, and eICU-CRD) and to harmonize concepts. The two cohorts differ in three ways: selection grain, expansion scope, and extraction method.

SCID is identified using source-specific ICD-9-CM and ICD-10-CM code lists developed with expert input. Because diagnostic ICDs are recorded at the hospital level, the selection grain for SCID is hospital admissions. The sepsis cohort, in contrast, is selected at the ICU-stay level using Sepsis-3 criteria [32], where suspected infection is identified from the timing of antibiotic administration and microbiology cultures, and qualifying stays have a Sequential Organ Failure Assessment (SOFA) score [35] of at least 2 within the corresponding infection window.

For SCID, rendehR expands extraction to patient context by default, anchored on the unit where the diagnosis is recorded. This preserves previous and subsequent admissions and ICU stays for the same patient, which may be clinically relevant even when the qualifying diagnosis is recorded during only one encounter.

In MIMIC-III, 4,320 hospital admissions satisfied the SCID definition, corresponding to 3,824 unique patients. Patient-level expansion retained all available records associated with these patients, resulting in 5,725 hospital admissions and 5,983 ICU stays in the cohort-specific dataset. In MIMIC-IV, 39,850 hospital admissions satisfied the SCID definition, corresponding to 26,177 unique patients. Expansion to patient context resulted in 104,988 hospital admissions and 17,405 ICU stays. In eICU-CRD, the SCID definition identified 2,603 qualifying ICU stays corresponding to 2,343 hospital admissions. After expansion to patient context, the retained patient table contained 2,823 hospital admissions of 2,259 unique patients with 3,441 ICU stay-level records associated with the selected patients.

Sepsis-3 is defined at the ICU-stay level in all three sources using the Sepsis-3 criteria, and by default it is not expanded to a higher level of granularity. The resulting cohorts contained 34460, 42296, and 47035 qualifying ICU stays in MIMIC-III, MIMIC-IV, and eICU-CRD, respectively. Sepsis cohort selection also differs from SCID in its method of extraction. SCID uses an R function to select qualifying records, while Sepsis-3 is implemented using source-specific SQL adapted from the MIT-LCP MIMIC-Code definitions [36].

### 3.2 Cohort restriction and data construction

Restricting source tables to the selected cohort substantially reduced the data retained for downstream concept construction particularly for rare conditions such as SCID. In the MIMIC-III SCID cohort, rendehR retained 34.9 million *chartevents* rows and 2.44 million *labevents* rows, compared with 330.71 and 27.85 rows in the corresponding source tables — a reduction of 90 % and 87 %, respectively. These reductions for the same tables are 81% and 88% for MIMIC-IV, respectively. For eICU-CRD, 4.21 million *nursecharting* rows and 3.98 million *vitalperiodic* rows, are kept, reductions of 97.2% relative to their source tables.

Across all cohort-filtered clinical tables, 10.79% of MIMIC-III source rows, 31.86% of MIMIC- IV and 2.69% of eICU-CRD were retained for the SCID cohort. For Sepsis-3, these numbers are 81.51%, 50.89%, and 38.15% for MIMIC-III, MIMIC-IV, and eICU-CRD, respectively, which is expected. Detailed per-table row counts and build times are provided in Supplementary Table 1- 3.

### 3.3 Clinical concept inventory

The current rendehR library contains 22 flat concepts, 123 time-series concepts, and 75 duration concepts. The atomic configuration includes 220 distinct concept names with source-specific mappings across MIMIC-III, MIMIC-IV, and eICU-CRD. Flat concepts include variables that are constant within a hospital admission, such as demographics, hospital and ICU length of stay, admission type, and admission location. Time-series concepts include repeated physiological and laboratory measurements such as heart rate, mean arterial pressure, creatinine, lactate, and glucose. Duration concepts represent interval-valued exposures and interventions.

The current duration library consists primarily of medication-exposure concepts adapted from BlendedICU [17]. These concepts harmonize exposure to a common active ingredient and its timing across sources. Route of administration, dose, formulation, and source-specific units may be retained where available but are not currently harmonized into a common representation. Additional duration concepts include selected non-medication interventions for which source- specific definitions are available.

The complete concept inventory is distributed with rendehR through the packaged concept configuration and source-specific SQL definitions and is also provided in Supplementary Table 4. Atomic mappings and curated concepts are available within the package, with curated concepts provided under the corresponding source-specific SQL directories.

Table 2 summarizes how the three frameworks’ concept vocabularies overlap; Supplementary Table 4 gives the concept-by-concept crosswalk. Of 170 distinct harmonized concepts across the three, only 51 (30%) are defined by all three. Ricu supports derived concepts such as the Horowitz (PaO₂/FiO₂) and AVPU (Alert, Verbal, Pain, Unresponsive) [37], which are not currently implemented in rendehR. Conversely, ricu in its current version does not provide concepts of ventilator settings such as positive end-expiratory pressure, plateau pressure, tidal volume and set respiratory rate and that is the reason no respiratory concept is shared by all three frameworks. BlendedICU is the narrowest, restricting itself to variables it can harmonize across five databases, and it is the only one to keep invasive and non-invasive blood pressure as separate concepts. rendehR is the broadest, and its surplus is concentrated in three places: analyte concepts absent from the other two such as anion gap, amylase, a specimen-aware design in which cerebrospinal, pleural, ascitic, urinary and stool analytes are modelled as concepts distinct from their blood counterparts where available. An injury-severity layer (e.g. injury severity score ISS, injury mechanism and intent) is also provided by rendehR as specific concepts related to SCID cohort.

**Table 1.**
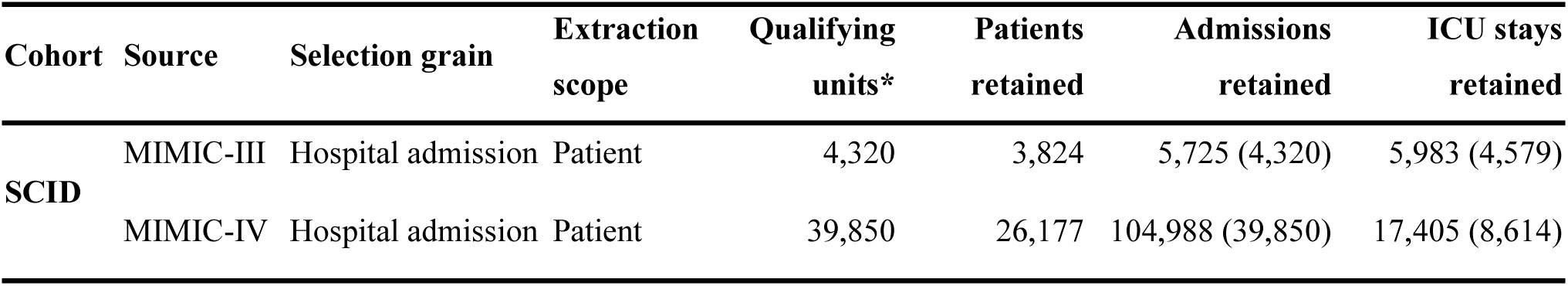
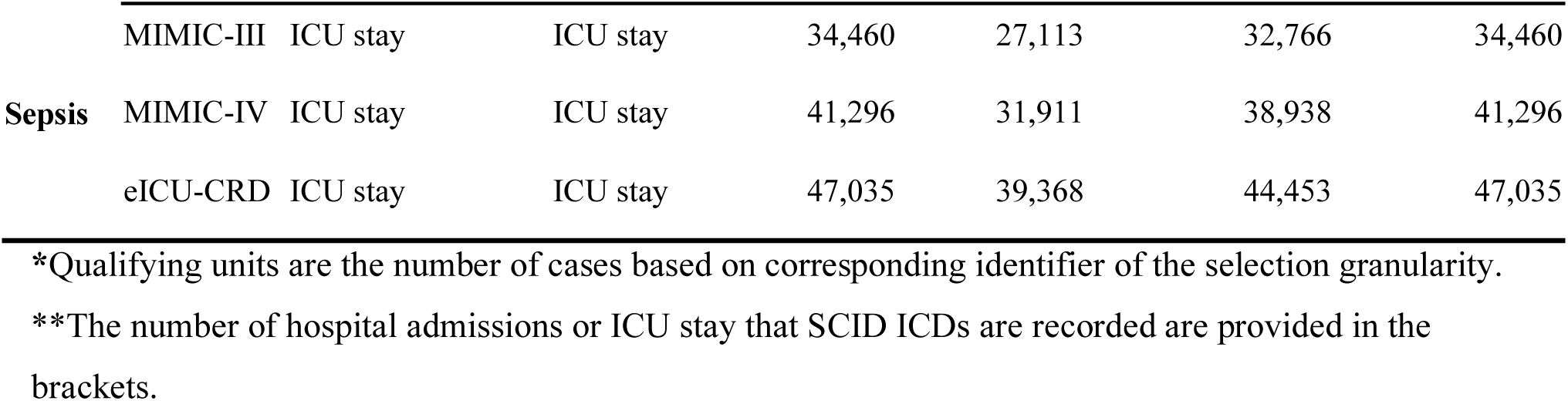
SCID and Sepsis-3 default cohort construction across sources.

| Cohort | Source | Selection grain | Extraction scope | Qualifying units* | Patients retained | Admissions retained | ICU stays retained |
|---|---|---|---|---|---|---|---|
| **SCID** | MIMIC-III | Hospital admission | Patient | 4,320 | 3,824 | 5,725 (4,320) | 5,983 (4,579) |
|  | MIMIC-IV | Hospital admission | Patient | 39,850 | 26,177 | 104,988 (39,850) | 17,405 (8,614) |
| **Sepsis** | MIMIC-III | ICU stay | ICU stay | 34,460 | 27,113 | 32,766 | 34,460 |
|  | MIMIC-IV | ICU stay | ICU stay | 41,296 | 31,911 | 38,938 | 41,296 |
|  | eICU-CRD | ICU stay | ICU stay | 47,035 | 39,368 | 44,453 | 47,035 |

\*Qualifying units are the number of cases based on corresponding identifier of the selection granularity.
\*\*The number of hospital admissions or ICU stay that SCID ICDs are recorded are provided in the brackets.

**Table 2.** Harmonized concept coverage by clinical domain. Counts are distinct harmonized concepts, not source-specific item identifiers: a concept is counted once per framework regardless of how many databases that framework maps it to. Shared is the number of concepts defined by all three frameworks.

| **Domain** | **rendehR** | **ricu** | **BlendedICU** | **Shared** |
|---|---|---|---|---|
| Vitals | 13 | 10 | 8 | 8 |
| Blood gas | 15 | 10 | 5 | 5 |
| Chemistry | 29 | 21 | 15 | 15 |
| Hematology | 26 | 20 | 4 | 4 |
| Respiratory | 8 | 6 | 6 | 0 |
| Neurological | 5 | 6 | 4 | 4 |
| Output | 1 | 2 | 1 | 1 |
| Medications | 8 | 12 | 10 | 8 |
| Demographics | 15 | 6 | 9 | 4 |
| Outcomes | 7 | 3 | 2 | 2 |
| Scores / derived | 21 | 8 | 0 | 0 |
| Injury severity | 7 | 0 | 0 | 0 |
| **Total** | **155** | **104** | **64** | **51** |

The comparison also identifies three concepts that ricu and BlendedICU both expose but rendehR currently derives only through database-specific SQL rather than as harmonized concepts: urine output, total Glasgow Coma Scale, and ventilation windows. Vasopressin exposure is absent from current version of rendehR and will be provided in the next version.

### 3.4 Validation

We validated rendehR in two ways: an internally written harness of 175 distinct checks that reuses no package code, and an external comparison against ricu v0.6.3. The harness was run first on demo datasets (1,345 checks) and then on the SCID cohort in the full datasets (302 checks), covering 46,513 admissions and 32,260 patients (2,343 eICU, 4,320 MIMIC-III, 39,850 MIMIC- IV) and 62.7 million harmonized time-series and duration rows. Raw counts were read from PhysioNet CSV files as untyped text, and concept selection, numeric parsing, unit conversion, ICD normalization, and cohort derivation were re-implemented from their specifications.

On full data, imported row counts matched the independent counts for all 88 tables, including chartevents at 34,891,373 rows in MIMIC-III and 82,669,177 in MIMIC-IV, with no rows rejected and no values lost in type conversion. Concept extraction reproduced the independent selection exactly for 136/198 concepts in MIMIC-III, 151/209 in MIMIC-IV, and 181/187 in eICU; each of the 126 shortfalls was confirmed row by row to be the documented chart-copy de-duplication. Sampled events were recomputed from raw offsets and raw value strings: 58,885 timestamps and 59,285 values, all in agreement. Age, sex, and in-hospital mortality matched a from-scratch recomputation across 113,536 admissions with 100% agreement in each source, and cohort membership matched the stored flag for all 46,513 admissions. The cohort artifact was also reproducible: identical whether built in one statement or in chunks, identical through the Parquet cache, and identical after an interrupted build was resumed; at full scale, two independently chunked builds of the complete SCID cohort produced 974 tables with zero differences. Table 3 provides the summary of the ten validation layers.

**Table 3.** Validation summary.

| Layer | Reference | Denominator (MIMIC-III/ MIMIC-IV/eICU-CRD) | Result |
|---|---|---|---|
| Imported table row counts | raw PhysioNet CSV files, read as untyped text | 88 tables, all sources | 100% agreed |
| Concept extraction | from-specification re-implementation (oracle) | 594 concept-source pairs (198 / 209 / 187) | 468 exact;<br>126 identical after deduplication of concepts* |
| Sampled timed events | raw offsets and value strings | 58,885 timestamps; 59,285 values | 100% agreed |
| Demographics | from-scratch recomputation | 113,536 admissions | 100% (age, sex, in-hospital mortality) |
| Cohort membership | stored cohort flag | 46,513 admissions | 100% agreed |
| Build determinism | independent single-statement / chunked rebuilds | 974 tables | 100% agreed |
| ricu: cohort | ricu v0.6.3, same raw files and admissions | 46,513 admissions (4,320 / 39,850 / 2,343) | Identical admissions, patients, and ICU stays |
| ricu: static variables | ricu v0.6.3 | 7 variables × 3 sources | 100% agreed for age, sex, mortality, hospital and ICU LOS;<br>84.6–100% for weight and height |
| ricu: time-series concepts | ricu v0.6.3, row for row on admission, minute, value | 193 concepts<br>(64 / 65 / 64) | 0 failures; 31 documented differences; first-value alignment 99.7–100%; KS distance mean 0.0010 (maximum 0.038, |
| ricu: exposures | ricu v0.6.3 | 7 concepts × 3 sources | all matched or documented (orders vs infusions; MIMIC-IV premixed phenylephrine 1,581 vs 1,294) |

The external comparison against ricu v0.6.3 used 80 matched concept pairs, covering demographics, static admission variables, laboratory and bedside time series, and vasoactive and insulin exposures, and compared both pipelines on exactly the same admissions. Cohort derivation agreed with the same cohort observations as in Table 1. Age, sex, in-hospital mortality, and hospital and ICU lengths of stay agreed for 100% of admissions. All mapped time-series concepts (64 in MIMIC-III, 65 in MIMIC-IV, 64 in eICU) reproduced ricu row for row on admission, minute, and value (193 comparisons, zero failures). First-value clock alignment, defined as the median per source of the proportion of admissions whose earliest concept measurement falls within ±1 minute, was 99.7–100%; representative values are MIMIC-IV heart rate 100.0% of 7,619 admissions, MIMIC-IV MAP 100%, MIMIC-III lactate 99.85%, and eICU creatinine 99.96%. Value distributions matched closely: across the 166 concepts compared directly, the mean Kolmogorov–Smirnov distance between rendehR and ricu was 0.0010 (range 0–0.038; 0.0010 in MIMIC-III, 0.0013 in MIMIC-IV and 0.0005 in eICU-CRD), 163 concepts had a distance of 0.01 or less, and none exceeded the 0.05 criterion. The largest distances were for SpO2 in MIMIC-III (0.038; median 98% in both packages), direct bilirubin in MIMIC-IV (0.023; median 1.0 vs 1.1 mg/dL) and magnesium in eICU-CRD (0.014; median 2.0 mg/dL in both). The 20 concepts with a registered definitional difference had a mean distance of 0.054 (range 0.002–0.145; largest for MIMIC-III troponin T, where rendehR also reads charted values), reflecting the different definitions (Supplementary Tables 6 and 7). Weight and height, where rendehR pools admit, daily and echo readings and ricu reads admit-weight items only, still agreed for 84.6–100% of admissions, and exposures matched wherever both packages read the same item sets.

Every remaining difference is registered with its measured magnitude and an item-level explanation. The registry contains 46 entries, including eICU exposures, where rendehR counts medication orders alongside infusions while ricu reads infusions only; ricu’s exclusion of venous pO₂ below its plausibility limit; and the MIMIC-IV phenylephrine exposure count (1,581 vs 1,294 admissions), traced to the premixed phenylephrine formulations (items 229630, 229632) that rendehR reads and ricu does not.

### 3.5 Downstream analysis

The current rendehR pipeline was applied to construct the SCID cohort. Supplementary Figure 1 provides an example of the harmonized physiological measurements, medication exposures, and flat variables obtained from one patient in this cohort, a 29-year-old female with traumatic SCI who was admitted through the emergency department in the MIMIC-IV demo dataset using Shiny app.

We also tested the harmonized output in a prediction task. We constructed an admission-level dataset from the MIMIC-III and MIMIC-IV SCID cohort to predict in-hospital mortality. The setup follows our previous work [38]. Age and sex were obtained from the flat tables. Twenty routine blood biomarker measurements recorded during the first 24 hours of admission were summarized using prespecified minimum, maximum, or mean values and joined to the flat table.

We fit a logistic regression model to predict in-hospital mortality using the R stats package [28]. Using an 70/30 training-test split, the model achieved a test-set area under the receiver operating characteristic curve (AUC) of 0.70 (95% CI, 0.64–0.76), consistent with the performance observed in our previous analysis using a similar prediction setting [38]. Figure 3 shows the ROC curve and estimated odds ratios with 95% confidence intervals for the model covariates.

**Figure 3:**
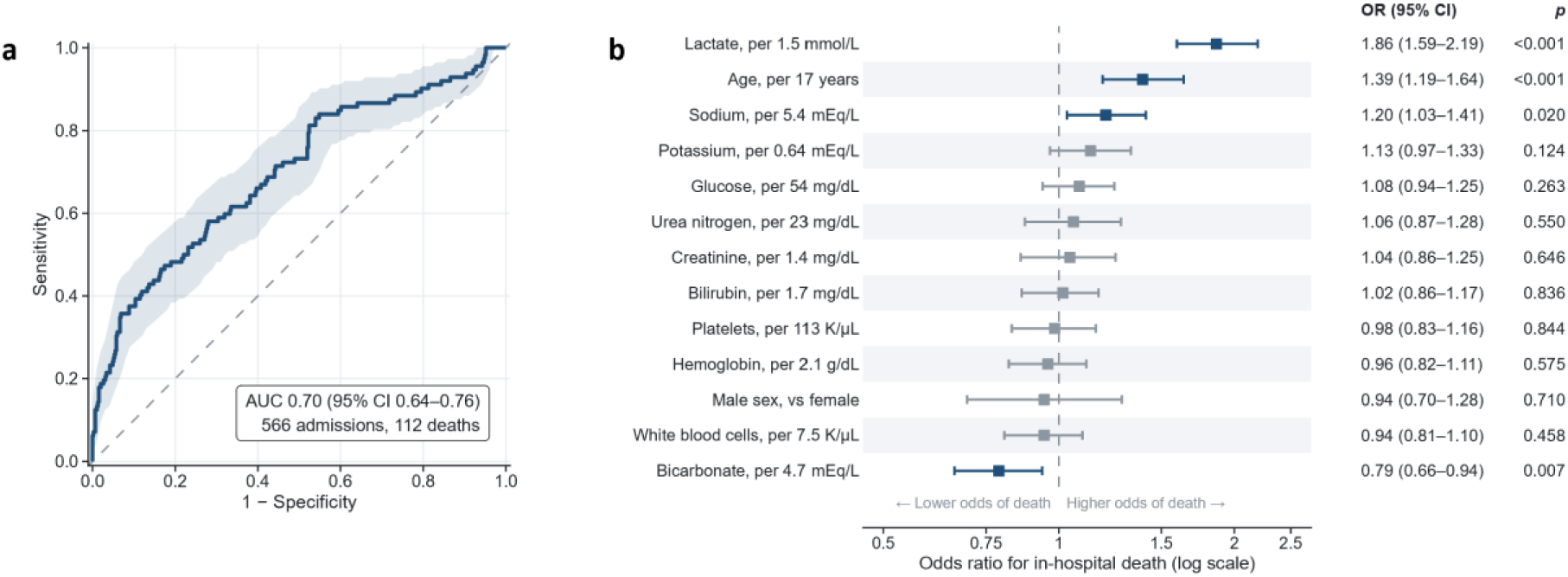
ROC curve (a) and estimated odds ratios with 95% confidence intervals for the model covariates (b) of in-hospital mortality prediction among SCID patients based on first day biomarkers.

The code of this analysis is provided in the Supplementary material.

## 4. Discussion

rendehR addresses a specific gap in existing multi-source EHR harmonization tools, which implement full database harmonization before any analysis. Many health researchers, however, are interested in the analysis of a defined cohort. Identifying the cohort first, rather than constructing a harmonized representation of all source data, substantially reduces the memory and computational power required. It also makes the workflow more scalable and makes adding sources more straightforward. In packages such as ricu or the BlendedICU pipeline, cohort construction can be implemented by the user, but only after going through harmonization of all data first. rendehR instead derives the study-specific dataset first and gives the user the option to keep intermediate outputs. This matters most for studies where the cohort is a small fraction of the source data, and where the researchers are interested in developing disease specific medical concepts.

Table 4 provides a comparison between similar existing packages and pipelines. Firstly, cohort identification in rendehR is implemented through key identifiers, either directly or indirectly after diagnostic ICD codes are chosen. At the cohort extraction stage, rendehR lets the user choose both the granularity of cohort definition and the scope of extraction. Separating selection grain from extraction scope allows cohort membership to follow the level at which the clinical definition is evaluated, while retaining broader linked context when required. The user can also define the unit of analysis to be hospital admission or the ICU stay. This differs from pipelines such as BlendedICU, where the main structure is based on ICU stays and users should do post hoc coding or analysis if they want to change this to admissions.

**Table 4.** rendehR relative to selected EHR data-processing frameworks.

| Tool | Primary purpose | Sources / setting | Cohort handling | Harmonization approach | Relationship to rendehR |
|---|---|---|---|---|---|
| MIMIC-Extract | ML-oriented preprocessing and representation | MIMIC-III | Task/population filtering within MIMIC-III | Standardized ML-ready variables and time series | Single-source and prediction-oriented, rendehR is multiple source |
| FIDDLE | Data-driven feature construction | MIMIC-III<br>eICU-CRD | Task-defined preprocessing | Feature-level preprocessing for prediction | Focuses on feature construction rather than persistent study databases |
| ricu | Unified ICU data access through clinical concepts | MIMIC-III<br>MIMIC-IV<br>eICU-CRD<br>HiRID<br>AmsterdamUMC | User selects observations/concepts from supported sources | Source-agnostic concepts with source-specific mappings | rendehR adopts the concept principle but executes and persists source-specific definitions as SQL in cohort-specific databases |
| BlendedICU | Cross-database ICU harmonization | MIMIC-III<br>MIMIC-IV<br>eICU-CRD<br>HiRID,<br>AmsterdamUMC | Produces a broad harmonized dataset | Common variables and OMOP-standardized representation | rendehR constructs study-specific cohorts and selected concepts rather than a fixed harmonized dataset |
| MIT-LCP | Reusable derived-variable SQL | MIMIC-III<br>MIMIC-IV<br>eICU-CRD | Not primarily a cohort-management framework | Curated source-specific SQL | rendehR adapts selected definitions and records their origin within its workflow |
| rcprd | Cohort-based extraction and analysis-ready data | CPRD EHR | Cohort defined before extraction | Database-backed variable extraction | A cohort-first analogue in primary care rather than heterogeneous public ICU databases |
| rendehR | Cohort-centered multi-source EHR dataset construction | MIMIC-III<br>MIMIC-IV<br>eICU-CRD | Executable cohort definition before cohort-specific materialization | Source-aware SQL concepts and harmonized outputs | Connects cohort definition, concept execution, persistent outputs, and provenance |

In ricu, the user can switch the ID structure while keeping the timing of the time window of events as specified, but rendehR gives the user more flexibility on how to choose the time window of merging the hospital-level events to a particular ICU when the granularity of the data changes from hospital level to ICU level. rendehR also gives researchers the ability to define the specific concepts that are most relevant to one field. The etiology of SCI table provided in the package, for example, is of value in studying an SCI cohort, but may be of less value to a researcher interested in coronary disease, who therefore does not have to build harmonized concepts related to it.

At the current version, rendehR has predefined cohorts of spinal cord injury and sepsis based on Sepsis-3 criteria. These cohorts could potentially be developed further to a range of conditions such as brain injuries or cardiovascular conditions, which would serve more researchers. Once the cohort is identified, the package filters all available data for each source and provides raw cohort files in Parquet form which are useful on its own, independent of the downstream concept layer. After a subset of the data is selected, medical concepts are created for a much smaller subset of the data relevant to the researcher. Like ricu, rendehR separates concept definitions from their execution, but executes them on SQL and persists those definitions in the concept tables. Both rendehR and ricu can extend their concepts. When the user is interested in the final output, rendehR, like ricu, can provide that, but the modification such as adding or modifying a concept, changing the refence timeline of measurements, and adding a new data source are relatively easier in rendehR’s pipeline with keeping the original sources and auditable SQLs, in comparison to call back functions defined in ricu. Such modifications in BlendedICU are done by changing the source code and pipeline directly.

### MCP and agent ready

rendehR can be operated by AI agents as well. Its MCP server is written in R, needs only jsonlite to start, and uses the installed rendehR for its database tools; it runs locally as a standard input/output server launched with Rscript, implements MCP protocol versions 2024-11-05 to 2025-06-18, and can therefore be connected to any MCP-compatible client, such as Claude Code, Codex, or Cursor. The client is configured with the path to the launcher shipped in the installed package, the R library that contains rendehR, and the project configuration file; credentials stay in that configuration and are never passed through the agent. Every tool is declared read-only: database tools open PostgreSQL sessions in read-only mode with a 30-second statement timeout, return aggregate counts and dictionary metadata rather than patient records, and cannot run pipeline stages or save concepts. The agent skills handle pipeline changes and produce reviewable R code that the user runs or authorizes. The server and the skills are tested with the package (the JSON-RPC handshake, argument validation, every tool, and the stdio transport), and the skills’ instructions are checked against the exported functions so that they follow the package as it evolves. This way, an agent can help a researcher locate source items, draft concept definitions, and inspect prepared cohorts, while every data-changing step remains explicit and auditable.

### Limitations and future work

rendehR currently depends on PostgreSQL for curated concept execution, harmonization, source merging, and most access functions. DuckDB already handles raw-source querying, cohort selection, cohort persistence, and selected concept execution. The full DuckDB backend support is planned for version 2. Source support is limited to MIMIC-III, MIMIC-IV, and eICU-CRD. Adding a new source is possible, but it still requires tasks such as source-specific identifier mappings, cohort logic, and concept definitions. Additionally, the duration concept library is currently dominated by medication exposures from the BlendedICU pipeline, where harmonization happens at the level of active ingredient and exposure timing rather than route, dose, formulation, or unit. While this is important information, further work is needed to harmonize units and doses of such medications.

When a cohort is identified based on ICDs, the cohort selection step uses few relatively small tables to identify qualified patients. However, for conditions such as sepsis, the sepsis criteria itself is a complex concept built from multiple atomic concepts. These concepts are relatively smaller than the whole concept set, but they still have to be harmonized in advance. Although the current structure of DuckDB and the use of SQL facilitate this process, it is still an extra step and involving larger tables makes it slower.

Future work targets three directions. Backend support will expand, including full DuckDB execution for users who prefer a lighter-weight workflow. New source databases and packaged reference cohorts will be added through the existing registration and cohort-definition framework. OMOP support and an event-log structure are also planned, which will let users adapt rendehR for research on event-log and process-mining analysis.

## 5. Conclusion

rendehR builds disease-specific datasets from heterogeneous EHR sources by keeping cohort definition and clinical concepts as explicit, executable parts of the pipeline. A study population, the clinical variables derived from it, and the final harmonized dataset stay traceable to one another. The current implementation supports MIMIC-III, MIMIC-IV, and eICU-CRD, with both diagnosis-based and SQL-defined cohort selection, but rendehR is built for scalability, and users can easily expand to new sources. Researchers using rendehR will spend less time repeating source-specific processing, and the datasets they produce are easier to inspect, reuse, and extend.

## Data Availability

Full MIMIC-III, MIMIC-IV, and eICU-CRD data are not redistributed directly in the rendehR package and must be obtained from PhysioNet under the corresponding access requirements. Demo workflows use the publicly available PhysioNet demo datasets.

https://physionet.org/

## Availability of Data and Software

rendehR is available at https://github.com/ATE-DRIVEN-lab/rendehR under the GNU General Public License, version 3 (GPL-3); version 1.0.0 is described here. Documentation, including the function reference and articles, is at https://ate-driven-lab.github.io/rendehR/. Full MIMIC-III, MIMIC-IV, and eICU-CRD data are not redistributed with the package and must be obtained separately from PhysioNet under the corresponding access requirements. Demo workflows use the packaged source configurations for the available PhysioNet demo datasets.

## Funding

This work was supported through awards by the Wings for Life Research Foundation to ATE and the Craig H. Neilsen Foundation to MMR.

## Competing Interests

The authors declare no competing interests.

## Author Contributions

MMR and VS contributed to conceptualization, methodology, software development, validation, formal analysis, and visualization. MMR prepared the original manuscript draft. ATE contributed to the conceptualization, methodology, validation, project administration, and supervision of the study. All authors contributed to the review and editing of the manuscript, read and approved the final version.

## Acknowledgements

We thank Parisa Razavi Yeganeh and Rahul Desai for their input during the early stages of this work. We acknowledge the developers and maintainers of ricu, whose source-aware concept framework informed the concept layer of rendehR. We also acknowledge the MIT Laboratory for Computational Physiology and the contributors to MIMIC, eICU-CRD, MIMIC-Code, and eICU- Code for developing and maintaining these research resources.

## AI Usage

Generative AI tools (including ChatGPT and Claude) were used to assist with language editing, improving manuscript readability, and supporting coding and software development. All AI- assisted suggestions and generated code were reviewed, verified, and modified as necessary by the authors to ensure accuracy, clarity, and consistency with the intended objectives. The authors take full responsibility for the final content of the manuscript and associated code.

## Notes

### Competing Interest Statement

The authors have declared no competing interest.

### Author Declarations

Office of research ethics of University of Waterloo gave ethical approval for the analysis in this work.

