## Supplementary Material for "rendehR: a cohort-first, provenance-aware, agentic-ready pipeline for harmonizing multi- source electronic health records"

### Supplementary materials

#### Supplementarily Figures

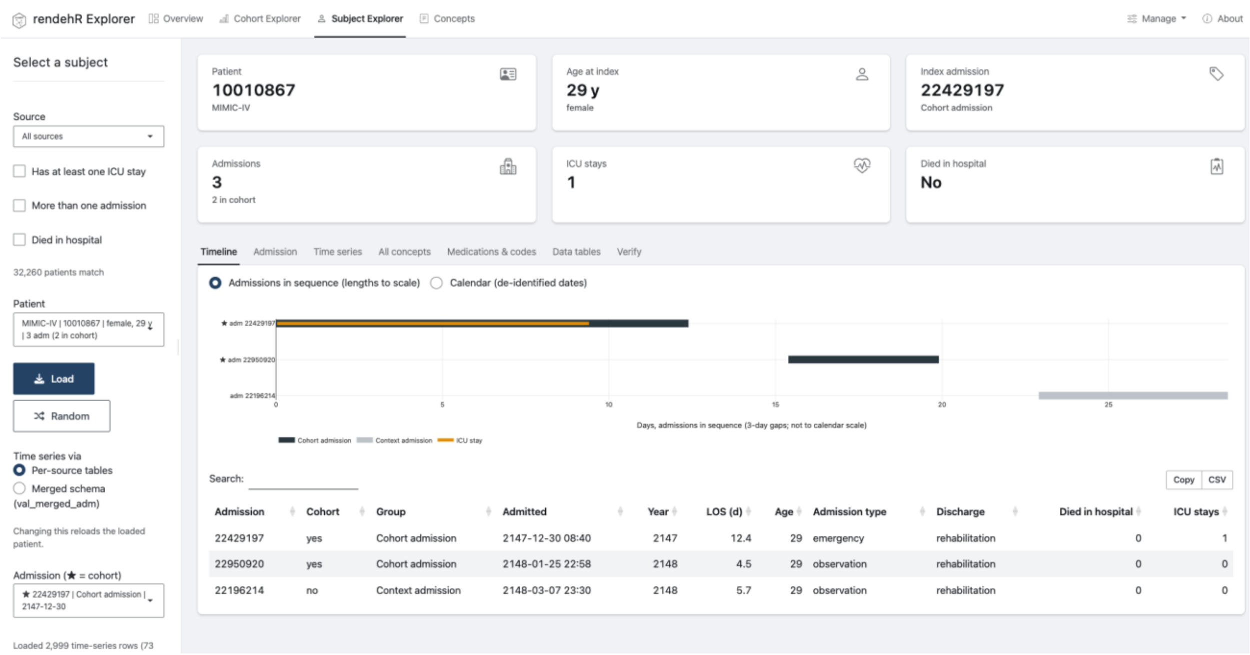


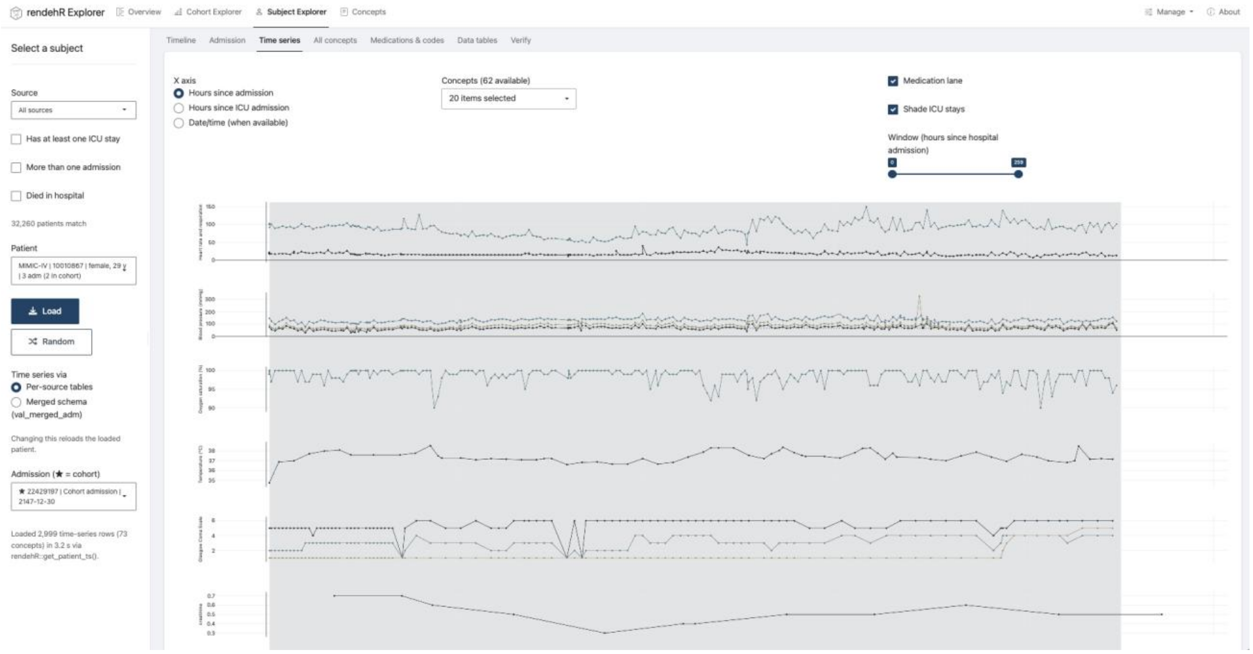

**Supplementary Figure 1:** Example of the rendehR Shiny app displaying demographic and admission-related information, including age, number of admissions, and length of stay (top panel), and time-series clinical concepts, including body temperature, blood pressure, and oxygen saturation (bottom panel), for a patient with spinal cord injury from MIMIC-IV demo.

#### Supplementary Tables

**Supplementary Table 1:** Row counts and coverage of cohorts for MIMIC-III

| | | SCID | | Sepsis-3 | |
|---|---|---|---|---|---|
| **Table Name** | **MIMIC-III (V)** | **Count** | **Coverage** | **Count** | **Coverage** |
| **admissions** | 58,976 | 5,725 | 9.71% | 32,766 | 55.56 |
| **callout** | 34,499 | 4,508 | 13.07% | 23,972 | 69.49 |
| **caregivers** | 7,567 | 7,567 | 100.00% | 7,567 | 100.00 |
| **chartevents** | 330,712,483 | 34,891,373 | 10.55% | 260,960,190 | 78.91 |
| **cptevents** | 573,146 | 79,115 | 13.80% | 463,838 | 80.93 |
| **d_cpt** | 134 | 134 | 100.00% | 134 | 100.00 |
| **d_icd_diagnoses** | 14,567 | 14,567 | 100.00% | 14,567 | 100.00 |
| **d_icd_procedures** | 3,882 | 3,882 | 100.00% | 3,882 | 100.00 |
| **d_items** | 12,487 | 12,487 | 100.00% | 12,487 | 100.00 |
| **d_labitems** | 753 | 753 | 100.00% | 753 | 100.00 |
| **datetimeevents** | 4,485,937 | 578,609 | 12.90% | 4,009,197 | 89.37 |
| **diagnoses_icd** | 651,047 | 79,472 | 12.21% | 428,850 | 65.87 |
| **drgcodes** | 125,557 | 13,682 | 10.90% | 74,617 | 59.43 |
| **icustays** | 61,532 | 5,983 | 9.72% | 34,460 | 56.00 |
| **inputevents_cv** | 17,527,935 | 1,381,428 | 7.88% | 13,736,540 | 78.37 |
| **inputevents_mv** | 3,618,991 | 511,422 | 14.13% | 3,187,929 | 88.09 |
| **labevents** | 27,854,055 | 2,441,888 | 8.77% | 16,730,484 | 60.06 |
| **microbiologyevents** | 631,726 | 81,396 | 12.88% | 501,708 | 79.42 |
| **noteevents** | 2,083,180 | 184,152 | 8.84% | 1,383,276 | 66.40 |
| **outputevents** | 4,349,218 | 495,412 | 11.39% | 3,486,710 | 80.17 |
| **patients** | 46,520 | 3,824 | 8.22% | 27,113 | 58.28 |
| **prescriptions** | 4,156,450 | 313,939 | 7.55% | 2,229,411 | 53.64 |
| **procedureevents_mv** | 258,066 | 36,361 | 14.09% | 207,190 | 80.29 |
| **procedures_icd** | 240,095 | 26,691 | 11.12% | 167,582 | 69.80 |
| **services** | 73,343 | 7,783 | 10.61% | 42,655 | 58.16 |
| **transfers** | 261,897 | 7,692 | 2.94% | 49,812 | 19.02 |

**Supplementary Table 2:** Row counts and coverage of cohorts for MIMIC-IV

| | | SCID | | Sepsis-3 | |
|---|---|---|---|---|---|
| **Table Name** | **MIMIC-IV (v3.1)** | **Count** | **Coverage** | **Count** | **Coverage** |
| **admissions** | 546,028 | 104,988 | 19.23 | 38,939 | 7.13 |
| **patients** | 364,627 | 26,177 | 7.18 | 31,911 | 8.75 |
| **transfers** | 2,413,581 | 393,733 | 16.31 | 211,702 | 8.77 |
| **d_hcpcs** | 89,208 | 89,208 | 100.00 | 89,208 | 100.00 |
| **diagnoses_icd** | 6,364,488 | 1,490,307 | 23.42 | 822,724 | 12.93 |
| **d_icd_diagnoses** | 112,107 | 112,107 | 100.00 | 112,107 | 100.00 |
| **d_icd_procedures** | 86,423 | 86,423 | 100.00 | 86,423 | 100.00 |
| **d_labitems** | 1,650 | 1,650 | 100.00 | 1,650 | 100.00 |
| **drgcodes** | 761,856 | 154,140 | 20.23 | 76,255 | 10.01 |
| **emar_detail** | 87,371,064 | 20,452,074 | 23.41 | 37,876,919 | 43.35 |
| **emar** | 42,808,593 | 9,646,960 | 22.54 | 10,879,064 | 25.41 |
| **hcpcsevents** | 186,074 | 31,174 | 16.75 | 89 | 0.05 |
| **labevents** | 158,374,764 | 17,697,248 | 11.17 | 26,101,113 | 16.48 |
| **microbiologyevents** | 3,988,224 | 394,978 | 9.90 | 638,301 | 16.00 |
| **pharmacy** | 17,847,567 | 3,904,078 | 21.87 | 4,259,462 | 23.87 |
| **poe_detail** | 8,504,982 | 1,756,158 | 20.65 | 2,964,455 | 34.85 |
| **poe** | 52,212,109 | 11,064,128 | 21.19 | 13,057,571 | 25.01 |
| **prescriptions** | 20,292,611 | 4,389,447 | 21.63 | 5,292,391 | 26.08 |
| **procedures_icd** | 859,655 | 164,577 | 19.14 | 197,372 | 22.96 |
| **services** | 593,071 | 116,012 | 19.56 | 51,383 | 8.66 |
| **chartevents** | 432,997,491 | 82,669,177 | 19.09 | 310,121,032 | 71.62 |
| **datetimeevents** | 9,979,761 | 1,823,087 | 18.27 | 7,646,195 | 76.62 |
| **d_items** | 4,095 | 4,095 | 100.00 | 4,095 | 100.00 |
| **icustays** | 94,458 | 17,405 | 18.43 | 41,296 | 43.72 |
| **inputevents** | 10,953,713 | 2,004,019 | 18.30 | 8,643,105 | 78.91 |

| Table Name | MIMIC-IV (v3.1) | SCID | | Sepsis-3 | |
|---|---|---|---|---|---|
| | | Count | Coverage | Count | Coverage |
| **outputevents** | 5,359,395 | 993,946 | 18.55 | 3,846,226 | 71.77 |
| **procedureevents** | 696,092 | 147,997 | 21.26 | 530,421 | 100.00 |
| **omr** | 7,753,027 | 1,423,353 | 18.36 | 1,258,892 | 16.24 |
| **caregiver** | 17,984 | 17,984 | 100.00 | 17,984 | 100.00 |
| **provider** | 42,244 | 42,244 | 100.00 | 42,244 | 100.00 |
| **ingredientevents** | 12,229,408 | 2,619,364 | 21.42 | 11,228,385 | 78.78 |

**Supplementary Table 3:** Row counts and coverage of cohorts for eICU-CRD

| Table Name | eICU (v2.0) | SCID | | Sepsis-3 | |
|---|---|---|---|---|---|
| | | Count | Coverage | Count | Coverage |
| **admissiondrug** | 874,920 | 41,313 | 4.72 | 384,357 | 43.93 |
| **admissiondx** | 626,858 | 10,395 | 1.66 | 161,708 | 25.80 |
| **allergy** | 251,949 | 8,328 | 3.31 | 93,513 | 37.12 |
| **apacheapsvar** | 171,177 | 3,006 | 1.76 | 43,716 | 25.54 |
| **apachepatientresult** | 297,064 | 5,318 | 1.79 | 79,952 | 26.91 |
| **apachepredvar** | 171,177 | 3,006 | 1.76 | 43,716 | 25.54 |
| **careplancareprovider** | 502,765 | 12,205 | 2.43 | 154,486 | 30.73 |
| **careplaneol** | 1,433 | 28 | 1.95 | 438 | 30.57 |
| **careplangeneral** | 3,115,018 | 76,794 | 2.47 | 979,583 | 31.45 |
| **careplangoal** | 504,139 | 18,712 | 3.71 | 180,850 | 35.87 |
| **careplaninfectiousdisease** | 8,056 | 240 | 2.98 | 2,481 | 30.80 |
| **customlab** | 1,082 | 10 | 0.92 | 424 | 39.19 |
| **diagnosis** | 2,710,672 | 135,301 | 4.99 | 1,149,924 | 42.42 |
| **hospital** | 208 | 208 | 100.00 | 208 | 100.00 |
| **infusiondrug** | 4,803,719 | 121,353 | 2.53 | 2,440,143 | 50.80 |
| **intakeoutput** | 12,030,289 | 373,097 | 3.10 | 6,114,905 | 50.83 |
| **lab** | 39,132,531 | 812,420 | 2.08 | 14,016,319 | 35.82 |
| **medication** | 7,301,853 | 135,421 | 1.85 | 3,145,196 | 43.07 |
| **microlab** | 16,996 | 791 | 4.65 | 3,369 | 19.82 |
| **note** | 2,254,179 | 66,666 | 2.96 | 722,726 | 32.06 |
| **nurseassessment** | 15,602,498 | 923,901 | 5.92 | 5,850,273 | 37.50 |

| Table Name | eICU (v2.0) | SCID | | Sepsis-3 | |
|---|---|---|---|---|---|
| | | Count | Coverage | Count | Coverage |
| **nursecare** | 8,311,132 | 451,963 | 5.44 | 2,734,524 | 32.90 |
| **nursecharting** | 151,604,232 | 4,205,962 | 2.77 | 66,544,486 | 43.89 |
| **pasthistory** | 1,149,180 | 27,167 | 2.36 | 413,571 | 35.99 |
| **patient** | 200,859 | 3,441 | 1.71 | 47,035 | 23.42 |
| **physicalexam** | 9,212,316 | 376,452 | 4.09 | 3,371,491 | 36.60 |
| **respiratorycare** | 865,381 | 42,324 | 4.89 | 432,292 | 49.95 |
| **respiratorycharting** | 20,168,176 | 804,782 | 3.99 | 10,607,421 | 52.60 |
| **treatment** | 3,688,745 | 165,590 | 4.49 | 1,191,307 | 32.29 |
| **vitalaperiodic** | 25,075,074 | 570,384 | 2.27 | 9,188,307 | 36.64 |
| **vitalperiodic** | 146,671,642 | 3,983,465 | 2.72 | 55,963,410 | 38.16 |

**Supplementary Table 4**: Full ricu, rendehR, and BlendedICU Concept Crosswalk

Legend: "—" denotes a concept the framework does not define. "†" denotes a concept the framework can produce only through database-specific derived SQL or an external mapping table, rather than as a source-agnostic harmonized concept; the responsible file is named in italics. Where a framework splits one concept into several, all names are listed. Superscripts refer to the notes following the table.

| Concept | ricu | rendehR | BlendedICU |
|---|---|---|---|
| **Vitals** | | | |
| Heart rate | hr | heart_rate | heart_rate |
| Systolic BP[1] | sbp | sbp | invasive_systolic_blood_pressure<br>noninvasive_systolic_blood_pressure |
| Diastolic BP | dbp | dbp | invasive_diastolic_blood_pressure<br>noninvasive_diastolic_blood_pressure |
| Mean arterial pressure | map | map | invasive_mean_blood_pressure<br>noninvasive_mean_blood_pressure |
| Temperature | temp | temperature | temperature |
| Respiratory rate | resp | resp_rate | respiratory_rate |
| $SpO_2$ (peripheral) | spo2 | spo2 | O2_pulseoxymetry_saturation |
| $SaO_2$ (arterial) | sao2 | so2 | O2_arterial_saturation |
| $O_2$ saturation (umbrella)[2] | o2sat | o2sat | — |

| Concept | ricu | rendehR | BlendedICU |
|---|---|---|---|
| End-tidal $CO_2$ | etco2 | etco2 | — |
| Intracranial pressure | — | icp | — |
| Pulmonary artery mean pressure | — | mpap | — |
| IABP mean pressure | — | iabp_mean | — |
| **Blood gas** | | | |
| pH | ph | ph | pH |
| $pO_2$ | po2 | po2 | paO2 |
| $pCO_2$ | pco2 | pco2 | paCO2 |
| Base excess | be | base_excess | — |
| Lactate | lact | lactate | lactate |
| $FiO_2$ | fio2 | fio2 | FiO2 |
| Ionized calcium | cai | calcium_ionized | — |
| Carboxyhemoglobin | hbco | hbco | — |
| Methemoglobin | methb | methemoglobin | — |
| Total $CO_2$ | tco2 | totalco2 | — |
| $HCO_3$ (calculated, blood gas)[3] | — | hco3 | — |
| Hematocrit (blood gas) | — | hematocrit_bloodgas | — |
| Hemoglobin (blood gas) | — | hemoglobin_bloodgas | — |
| Sodium (blood gas) | — | sodium_bloodgas | — |
| Potassium (blood gas) | — | potassium_bloodgas | — |
| **Chemistry** | | | |
| Albumin | alb | albumin | albumin |
| Alkaline phosphatase | alp | alp | alkaline_phosphatase |
| ALT | alt | alt | alanine_aminotransferase |
| AST | ast | ast | aspartate_aminotransferase |
| Bilirubin, total | bili | bili | bilirubine |
| Bilirubin, direct | bili_dir | bili_dir | — |
| Bilirubin, indirect | — | bili_indirect | — |
| BUN | bun | bun | blood_urea_nitrogen |
| Calcium, total | ca | calcium_total | calcium |
| Chloride | cl | chloride | chloride |
| Creatinine | crea | creatinine | creatinine |
| Glucose | glu | glucose | blood_glucose |
| Sodium | na | sodium | sodium |
| Potassium | k | potassium | potassium |
| Magnesium | mg | magnesium | magnesium |
| Phosphate | phos | phosphate | phosphate |
| Bicarbonate (lab) | bicar | bicarbonate | bicarbonate |
| C-reactive protein | crp | c_reactive_protein | — |

| Concept | ricu | rendehR | BlendedICU |
|---|---|---|---|
| Creatine kinase | ck | creatine_kinase | — |
| Creatine kinase MB | ckmb | creatine_kinase_MB | — |
| CK-MB index | — | creatine_kinase_mb_index | — |
| Troponin T | tnt | troponin_t | — |
| Troponin I | tri | troponin_i | — |
| Anion gap | — | anion_gap | — |
| Amylase | — | amylase | — |
| GGT | — | ggt | — |
| LDH | — | ldh | — |
| NT-proBNP | — | ntprobnp | — |
| Glucose, fingerstick/POC | — | glucose_fingerstick | — |
| **Hematology** | | | |
| White blood cells | wbc | wbc | white_blood_cells |
| Hemoglobin | hgb | hemoglobin | hemoglobin |
| Hematocrit | hct | hematocrit | — |
| Platelets | plt | platelet | platelets |
| Red blood cells | rbc | rbc | — |
| MCV | mcv | mcv | — |
| MCH | mch | mch | — |
| MCHC | mchc | mchc | — |
| RDW | rdw | rdw | — |
| Neutrophils % | neut | neutrophils | — |
| Neutrophils absolute | — | neutrophils_abs | — |
| Lymphocytes % | lymph | lymphocytes | — |
| Lymphocytes absolute | — | lymphocytes_abs | — |
| Monocytes % | — | monocytes | — |
| Monocytes absolute | — | monocytes_abs | — |
| Eosinophils % | eos | eosinophils | — |
| Eosinophils absolute | — | eosinophils_abs | — |
| Basophils | basos | basos | — |
| Band neutrophils | bnd | band | — |
| Prothrombin time | pt | pt | — |
| PTT | ptt | ptt | PTT |
| INR | inr_pt | prothrombin_time_inr | — |
| Fibrinogen | fgn | fibrinogen | — |
| D-dimer | — | d_dimer | — |
| ESR | esr | erythrocyte_sedimentation_rate | — |
| HbA1c | hba1c | hba1c | — |
| **Respiratory** | | | |
| PEEP[4] | — | peep | PEEP |

| Concept | ricu | rendehR | BlendedICU |
|---|---|---|---|
| Plateau pressure | — | plateau_pressure | plateau_pressure |
| Tidal volume, set | — | tidal_volume_setting | tidal_volume_setting |
| Tidal volume, expired | — | expiratory_tidal_volume | expiratory_tidal_volume |
| Tidal volume, spontaneous | — | tidal_volume_spontaneous | — |
| Respiratory rate, set | — | resp_rate_setting | respiratory_rate_setting |
| Ventilator mode | mech_vent | — | ventilator_mode |
| Ventilation start / end / windows | vent_startvent_endvent_ind | † ventilation.sql | — |
| Tracheostomy / intubated-GCS flag | ett_gcs | — | — |
| Supplemental oxygen | supp_o2 | — | — |
| P/F ratio (Horowitz) | pafi | † first_day_sofa.sql | — |
| S/F ratio | safi | — | — |
| **Neurological** | | | |
| GCS eye[5] | egcs | egcs | glasgow_coma_score_eye |
| GCS motor | mgcs | mgcs | glasgow_coma_score_motor |
| GCS verbal | vgcs | vgcs | glasgow_coma_score_verbal |
| GCS total[6] | tgcsgcs | † gcs.sql | glasgow_coma_score |
| RASS | rass | rass | — |
| AVPU | avpu | — | — |
| **Output** | | | |
| Urine output[7] | urine | † urine_output.sql | urine_output |
| Urine output per 24h | urine24 | — | — |
| **Medications** | | | |
| Antibiotics | abx | antibiotics | † ingredient mapping |
| Norepinephrine rate | norepi_rate | † norepinephrine_exp | † ingredient mapping |
| Epinephrine rate | epi_rate | † epinephrine_exp | † ingredient mapping |
| Dopamine rate | dopa_rate | † dopamine_exp | † ingredient mapping |
| Dobutamine rate | dobu_rate | † dobutamine_exp | † ingredient mapping |
| Phenylephrine rate | phn_rate | † phenylephrine_exp | † ingredient mapping |
| Vasopressin rate[8] | adh_rate | — | † ingredient mapping |
| Insulin | ins | † insulin_exp | † ingredient mapping |
| Dextrose | dex | † dextrose_exp | † ingredient mapping |
| Corticosteroids | cort | — | † ingredient mapping |
| Vasopressor durations (per drug) | norepi_durepi_dur<br>dopa_durdobu_dur | † **_exp (105 drugs)* | — |
| Norepinephrine equivalents | norepi_equiv | — | — |
| Vasopressor indicator | vaso_ind | — | — |

| Concept | ricu | rendehR | BlendedICU |
|---|---|---|---|
| Drug exposure library (breadth)[10] | 17 medication concepts | 105 *_exp concepts (78 with SQL) | 107 ingredients / 5k product map |
| **Demographics** | | | |
| Age | age | age | age |
| Sex | sex | sex | sex |
| Height | height | height | raw_height |
| Weight[9] | weight | weight_admit | raw_weight |
| BMI | bmi | — | — |
| Race / ethnicity | — | race | — |
| Insurance | — | insurance | — |
| Language | — | language | — |
| Marital status | — | marital_status | — |
| Care site | — | care_site | care_site |
| Admission type | adm | admission_type | — |
| Admission service | — | admission_service | — |
| Hospital admission location / origin | — | hospital_admission_location | origin |
| Hospital discharge location | — | hospital_discharge_location | discharge_location |
| ICU unit type (first) | — | icu_first_careunit | unit_type |
| ICU unit type (last) | — | icu_last_careunit | — |
| US vs non-US source flag | — | — | from_US |
| **Outcomes** | | | |
| Hospital mortality | death | hospital_mortality | mortality |
| ICU mortality | — | icu_mortality | — |
| Hospital LOS | los_hosp | hospital_los | — |
| ICU LOS | los_icu | icu_los | lengthofstay |
| 30-day readmission | — | readmission_30d | — |
| Time to death from admission | — | time_to_death_from_admission | — |
| Time to death from discharge | — | time_to_death_from_discharge | — |
| **Scores / derived** | | | |
| SOFA (+6 components)[11] | sofa + sofa_resp/coag/liver/cardio/cns/renal | † sofa.sql | — |
| qSOFA | qsofa | † qsofa.sql | — |
| SIRS | sirs | † sirs.sql | — |
| Sepsis-3 | sep3 | † sepsis3.sql | — |
| Suspected infection | susp_inf | † suspicion_of_infection.sql | — |
| MEWS | mews | — | — |
| NEWS | news | — | — |

| Concept | ricu | rendehR | BlendedICU |
|---|---|---|---|
| APS III | — | † apsiii.sql | — |
| SAPS II | — | † sapsii.sql | — |
| OASIS | — | † oasis.sql | — |
| LODS | — | † lods.sql | — |
| Charlson comorbidity index | — | † charlson.sql | — |
| Elixhauser comorbidity (AHRQ + Quan) | — | † elixhauser_ahrq_v37.sql / elixhauser_quan.sql / scores | — |
| Sepsis, pre-Sepsis-3 definitions | — | † angus.sql / martin.sql / explicit.sql | — |
| SAPS (original) / mLODS | — | † saps.sql / mlods.sql | — |
| CCS diagnosis grouping | — | † ccs_dx.sql | — |
| Code status | — | † code_status.sql | — |
| Arterial / central line durations | — | † arterial_line_durations.sql / central_line_durations.sql | — |
| KDIGO AKI stage | — | † kdigo_stages.sql | — |
| MELD | — | † meld.sql | — |
| RRT / CRRT | — | † rrt.sql + crrt.sql | — |
| Invasive line | — | † invasive_line.sql | — |
| Echo report data | — | echo_data | — |
| Body fluid sampling | samp | — | — |
| Diagnoses / comorbidity mapping | — | ICD SCI + injury-cause filters | 40-group diagnosis map |
| **Injury severity** | | | |
| Injury Severity Score (ISS)[13] | — | iss | — |
| New ISS (NISS) | — | niss | — |
| Max AIS | — | maxais | — |
| ISS computability status | — | iss_status | — |
| Injury mechanism | — | injury_mechanism | — |
| Injury intent | — | injury_intent | — |
| Injury cause status | — | injury_cause_status | — |
| **Specimen variants** | | | |
| Non-blood specimen analytes (37 concepts)[12] | — | † 37 concepts suffixed by specimen | — |

1. **BlendedICU** models invasive and non-invasive blood pressure as separate concepts; **ricu** and **rendehR** merge them. The same applies to diastolic and mean pressure.
2. A **rendehR** union of the peripheral and arterial concepts; **ricu**'s similarly named concept is a distinct respiratory concept.
3. **rendehR** separates blood-gas bicarbonate from the chemistry-panel measurement; the other two frameworks have one bicarbonate concept.
4. Ventilator settings are absent from **ricu** entirely.

5. **BlendedICU** maps the three Glasgow Coma Scale components for all databases except eICU.
6. **rendehR** provides the three components as harmonized concepts but derives the total only in MIMIC-specific SQL.
7. **rendehR**'s *_*urine* concepts are urinary *analytes*; urine volume is produced only by derived SQL.
8. Not represented in **rendehR** in any layer.
9. **ricu** treats weight as a time series; the other two take a single admission value.
10. **ricu** emits infusion rates for a small set of vasoactive drugs, **rendehR** emits dose-and-duration exposure intervals for 105 drugs (78 with generated SQL; the remainder are eICU-only configuration rows), and **BlendedICU** maps drug products onto 107 ingredients.
11. **ricu** computes SOFA and its components from other concepts; **rendehR** ships per-database SQL.
12. A **rendehR** design choice: cerebrospinal, pleural, ascitic, joint-fluid, urinary and stool analytes are modelled as concepts distinct from their blood counterparts, so the blood concept stays unambiguous. Neither comparator models specimen type.
13. The injury-severity layer is specific to **rendehR**'s spinal-cord-injury focus and is available for all three databases.

**Supplementary Table 5:** Comparison of running time for each stage using 16GB RAM.

| Cohort | Process | Source | Time (Second) |
|---|---|---|---|
| Sepsis | Import cohort | MIMIC-III | 829.0s |
| | Run DB | MIMIC-III | |
| | Run concepts | MIMIC-III | 12221.9s |
| Sepsis | Import cohort | MIMIC-IV | |
| | Run DB | MIMIC-IV | |
| | Run concepts | MIMIC-IV | |
| Sepsis | Import cohort | eICU-CRD | 401.0 |
| | Run DB | eICU-CRD | 2521.5s |
| | Run concepts | eICU-CRD | |
| Sepsis | Merging | | |
| SCID | Import cohort | MIMIC-III | 2236.4s* |
| | Run DB | MIMIC-III | 1533.3s |
| | Run concepts | MIMIC-III | 736.9s |
| SCID | Import cohort | MIMIC-IV | 580.5s |
| | Run DB | MIMIC-IV | 6535.9s |
| | Run concepts | MIMIC-IV | 2462.8s |
| SCID | Import cohort | eICU-CRD | 68.1s |
| | Run DB | eICU-CRD | 159.5s |
| | Run concepts | eICU-CRD | 887.0s |

*Cohort is generated from the CSV.GZ files directly.

**Supplementary Table 6:** KS Summary

| Data | Cohort | Concepts | Source | n | Mean D | Median D | Range | Largest D (concept) | D <= 0.01 | D > 0.05 |
|---|---|---|---|---|---|---|---|---|---|---|
| Full | SCID | compared directly | eICU-CRD | 54 | 0.0005 | 0 | 0.000-0.014 | 0.014 (magnesium) | 53 | 0 |
| Full | SCID | compared directly | MIMIC-III | 54 | 0.001 | 0.0002 | 0.000-0.038 | 0.038 (spo2) | 53 | 0 |
| Full | SCID | compared directly | MIMIC-IV | 58 | 0.0013 | 0.0004 | 0.000-0.023 | 0.023 (bili_dir) | 57 | 0 |
| Full | SCID | compared directly | all | 166 | 0.001 | 0.0001 | 0.000-0.038 | 0.038 (spo2) | 163 | 0 |
| Full | SCID | registered definitional difference | all | 20 | 0.0538 | 0.0387 | 0.002-0.145 | 0.145 (troponin_t) | 5 | 8 |
| Demo | General | compared directly | eICU-CRD | 54 | 0.0009 | 0 | 0.000-0.027 | 0.027 (magnesium) | 53 | 0 |
| Demo | General | compared directly | MIMIC-III | 54 | 0.0006 | 0 | 0.000-0.007 | 0.007 (hemoglobin) | 54 | 0 |
| Demo | General | compared directly | MIMIC-IV | 57 | 0.0018 | 0.0006 | 0.000-0.025 | 0.025 (creatine_kinase_mb) | 54 | 0 |
| Demo | General | compared directly | all | 165 | 0.0011 | 0 | 0.000-0.027 | 0.027 (magnesium) | 161 | 0 |
| Demo | General | registered definitional difference | all | 20 | 0.0658 | 0.0477 | 0.002-0.294 | 0.294 (troponin_t) | 4 | 9 |
| Demo | SCID | compared directly | eICU-CRD | 54 | 0.0004 | 0 | 0.000-0.009 | 0.009 (magnesium) | 54 | 0 |
| Demo | SCID | compared directly | MIMIC-III | 54 | 0.0024 | 0 | 0.000-0.045 | 0.045 (troponin_t) | 48 | 0 |
| Demo | SCID | compared directly | MIMIC-IV | 53 | 0.0001 | 0 | 0.000-0.006 | 0.006 (glucose) | 53 | 0 |
| Demo | SCID | compared directly | all | 161 | 0.001 | 0 | 0.000-0.045 | 0.045 (troponin_t) | 155 | 0 |
| Demo | SCID | registered definitional difference | all | 17 | 0.0994 | 0.075 | 0.004-0.348 | 0.348 (calcium_ionized) | 2 | 12 |

**Supplementary Table 7:** KS per concept

| Source | rendehR concept | ricu concept | Values rendehR | Values ricu | Median rendehR | Median ricu | KS D | p-value | Comparison |
|---|---|---|---|---|---|---|---|---|---|

| | | | | | | | | |
|---|---|---|---|---|---|---|---|---|
| eICU-CRD albumin | alb | 6,883 | 6,883 | 2.6 | 2.6 | 0 | 1 | compared |
| eICU-CRD alp | alp | 5,979 | 5,979 | 85 | 85 | 0 | 1 | compared |
| eICU-CRD alt | alt | 6,065 | 6,065 | 33 | 33 | 0 | 1 | compared |
| eICU-CRD ast | ast | 6,164 | 6,164 | 34 | 34 | 0 | 1 | compared |
| eICU-CRD band | bnd | 958 | 958 | 5 | 5 | 0 | 1 | compared |
| eICU-CRD base_excess | be | 6,440 | 6,440 | 0.7 | 0.7 | 0 | 1 | compared |
| eICU-CRD basos | basos | 10,991 | 10,991 | 0 | 0 | 0 | 1 | compared |
| eICU-CRD bicarbonate | bicar | 22,428 | 22,428 | 26 | 26 | 0 | 1 | compared |
| eICU-CRD bili | bili | 5,965 | 5,965 | 0.6 | 0.6 | 0 | 1 | compared |
| eICU-CRD bili_dir | bili_dir | 944 | 944 | 0.21 | 0.21 | 0 | 1 | compared |
| eICU-CRD bun | bun | 22,832 | 22,832 | 18 | 18 | 0 | 1 | compared |
| eICU-CRD c_reactive_protein | crp | 299 | 299 | 120 | 120 | 0 | 1 | compared |
| eICU-CRD calcium_ionized | cai | 2,061 | 847 | 1.088 | 1.15 | 0.0568 | 0.0418 | registered definitional difference |
| eICU-CRD calcium_total | ca | 22,273 | 22,273 | 8.4 | 8.4 | 0 | 1 | compared |
| eICU-CRD chloride | cl | 22,841 | 22,841 | 104 | 104 | 0 | 1 | compared |
| eICU-CRD creatine_kinase | ck | 1,448 | 1,448 | 426.5 | 426.5 | 0 | 1 | compared |
| eICU-CRD creatine_kinase_mb | ckmb | 679 | 679 | 4.1 | 4.1 | 0 | 1 | compared |
| eICU-CRD creatinine | crea | 22,899 | 22,899 | 0.76 | 0.76 | 0 | 1 | compared |
| eICU-CRD dbp | dbp | 1,177,482 | 705,158 | 63 | 61 | 0.0404 | <0.0001 | registered definitional difference |
| eICU-CRD egcs | egcs | 93,917 | 93,917 | 4 | 4 | 0 | 1 | compared |
| eICU-CRD eosinophils | eos | 11,649 | 11,673 | 1 | 1 | 0.0002 | 1 | compared |
| eICU-CRD fibrinogen | fgn | 565 | 565 | 296 | 296 | 0 | 1 | compared |
| eICU-CRD fio2 | fio2 | 115,133 | 114,583 | 40 | 40 | 0.0009 | 1 | compared |
| eICU-CRD glucose | glu | 23,275 | 74,371 | 122 | 131 | 0.1039 | <0.0001 | registered definitional difference |
| eICU-CRD hbco | hbco | 1,672 | 1,672 | 0.9 | 0.9 | 0 | 1 | compared |
| eICU-CRD heart_rate | hr | 3,653,003 | 3,285,931 | 86 | 86 | 0.0019 | <0.0001 | registered definitional difference |
| eICU-CRD hematocrit | hct | 23,606 | 23,606 | 29.7 | 29.7 | 0 | 1 | compared |
| eICU-CRD hemoglobin | hgb | 23,775 | 23,775 | 9.7 | 9.7 | 0 | 1 | compared |
| eICU-CRD lactate | lact | 3,305 | 3,305 | 1.6 | 1.6 | 0 | 1 | compared |
| eICU-CRD lymphocytes | lymph | 11,865 | 11,877 | 11.5 | 11.5 | 0.0003 | 1 | compared |
| eICU-CRD magnesium | mg | 12,327 | 12,326 | 2 | 2 | 0.014 | 0.1759 | compared |

| | | | | | | | | | |
|---|---|---|---|---|---|---|---|---|---|
| eICU-CRD | map | map | 1,264,956 | 1,182,955 | 81 | 81 | 0.0016 | 0.0895 | registered definitional difference |
| eICU-CRD | mch | mch | 19,658 | 19,658 | 29.9 | 29.9 | 0 | 1 | compared |
| eICU-CRD | mchc | mchc | 21,114 | 21,114 | 33 | 33 | 0 | 1 | compared |
| eICU-CRD | mcv | mcv | 21,128 | 21,128 | 90 | 90 | 0 | 1 | compared |
| eICU-CRD | methemoglobin | methb | 1,633 | 1,633 | 0.5 | 0.5 | 0 | 1 | compared |
| eICU-CRD | mgcs | mgcs | 93,876 | 93,876 | 6 | 6 | 0 | 1 | compared |
| eICU-CRD | neutrophils | neut | 10,639 | 10,639 | 77.5 | 77.5 | 0 | 1 | compared |
| eICU-CRD | o2sat | o2sat | 3,211,399 | 3,211,399 | 98 | 98 | 0 | 1 | compared |
| eICU-CRD | pco2 | pco2 | 7,649 | 7,649 | 40.9 | 40.9 | 0 | 1 | compared |
| eICU-CRD | ph | ph | 7,625 | 7,625 | 7.39 | 7.39 | 0 | 1 | compared |
| eICU-CRD | phosphate | phos | 9,623 | 9,623 | 3.2 | 3.2 | 0 | 1 | compared |
| eICU-CRD | platelet | plt | 21,781 | 21,781 | 212 | 212 | 0 | 1 | compared |
| eICU-CRD | po2 | po2 | 7,533 | 7,533 | 97 | 98 | 0 | 1 | compared |
| eICU-CRD | potassium | k | 25,411 | 25,411 | 4 | 4 | 0 | 1 | compared |
| eICU-CRD | prothrombin_time_inr | inr_pt | 6,455 | 6,455 | 1.2 | 1.2 | 0 | 1 | compared |
| eICU-CRD | pt | pt | 6,211 | 6,211 | 14.7 | 14.7 | 0 | 1 | compared |
| eICU-CRD | ptt | ptt | 4,586 | 4,586 | 32 | 32 | 0 | 1 | compared |
| eICU-CRD | rass | rass | 45,934 | 45,934 | 0 | 0 | 0 | 1 | compared |
| eICU-CRD | rbc | rbc | 21,505 | 21,505 | 3.33 | 3.33 | 0 | 1 | compared |
| eICU-CRD | rdw | rdw | 20,343 | 20,343 | 14.9 | 14.9 | 0 | 1 | compared |
| eICU-CRD | resp_rate | resp | 3,211,292 | 2,858,394 | 18 | 18 | 0.0038 | <0.0001 | registered definitional difference |
| eICU-CRD | sbp | sbp | 1,178,004 | 705,693 | 126 | 128 | 0.0392 | <0.0001 | registered definitional difference |
| eICU-CRD | so2 | sao2 | 5,912 | 5,912 | 97 | 97 | 0 | 1 | compared |
| eICU-CRD | sodium | na | 24,952 | 24,952 | 139 | 139 | 0 | 1 | compared |
| eICU-CRD | spo2 | spo2 | 3,205,487 | 3,205,487 | 98 | 98 | 0 | 1 | compared |
| eICU-CRD | temperature | temp | 405,241 | 391,888 | 37.44 | 37.44 | 0.0028 | 0.0936 | compared |
| eICU-CRD | totalco2 | tco2 | 2,274 | 2,324 | 25 | 25 | 0.0094 | 1 | compared |
| eICU-CRD | troponin_t | tnt | 206 | 206 | 0.06 | 0.06 | 0 | 1 | compared |
| eICU-CRD | vgcs | vgcs | 92,464 | 92,464 | 4 | 4 | 0 | 1 | compared |
| eICU-CRD | wbc | wbc | 21,589 | 21,589 | 10.4 | 10.4 | 0 | 1 | compared |
| MIMIC-III | albumin | alb | 6,836 | 6,830 | 2.9 | 2.9 | 0.0004 | 1 | compared |
| MIMIC-III | alp | alp | 9,448 | 9,436 | 108 | 108 | 0.0008 | 1 | compared |
| MIMIC-III | alt | alt | 9,692 | 9,679 | 34 | 34 | 0.0005 | 1 | compared |
| MIMIC-III | ast | ast | 9,682 | 9,670 | 39 | 39 | 0.0004 | 1 | compared |

| | | | | | | | | | |
|---|---|---|---|---|---|---|---|---|---|
| MIMIC-III | band | bnd | 3,530 | 3,525 | 1 | 1 | 0.0005 | 1 | compared |
| MIMIC-III | base_excess | be | 39,103 | 38,971 | 0 | 0 | 0.0002 | 1 | compared |
| MIMIC-III | basos | basos | 8,416 | 8,409 | 0.2 | 0.2 | 0.0002 | 1 | compared |
| MIMIC-III | bicarbonate | bicar | 53,734 | 53,687 | 26 | 26 | 0.0002 | 1 | compared |
| MIMIC-III | bili | bili | 9,888 | 9,855 | 0.7 | 0.7 | 0.0009 | 1 | compared |
| MIMIC-III | bili_dir | bili_dir | 794 | 793 | 1 | 1 | 0.0009 | 1 | compared |
| MIMIC-III | bun | bun | 54,936 | 54,814 | 19 | 19 | 0.0002 | 1 | compared |
| MIMIC-III | c_reactive_protein | crp | 588 | 587 | 79.6 | 79.6 | 0.0017 | 1 | compared |
| MIMIC-III | calcium_ionized | cai | 21,136 | 20,864 | 1.12 | 1.12 | 0.0022 | 1 | compared |
| MIMIC-III | calcium_total | ca | 45,434 | 45,355 | 8.4 | 8.4 | 0.0003 | 1 | compared |
| MIMIC-III | chloride | cl | 60,432 | 54,844 | 104 | 104 | 0.012 | 0.0005 | registered definitional difference |
| MIMIC-III | creatine_kinase | ck | 7,297 | 7,277 | 154 | 154 | 0.0006 | 1 | compared |
| MIMIC-III | creatine_kinase_mb | ckmb | 4,908 | 4,896 | 5 | 5 | 0.0006 | 1 | compared |
| MIMIC-III | creatinine | crea | 55,053 | 54,930 | 0.8 | 0.8 | 0.0002 | 1 | compared |
| MIMIC-III | dbp | dbp | 556,750 | 558,081 | 61 | 61 | 0.0001 | 1 | compared |
| MIMIC-III | egcs | egcs | 160,562 | 160,628 | 4 | 4 | 0.0001 | 1 | compared |
| MIMIC-III | eosinophils | eos | 8,411 | 8,407 | 0.8 | 0.8 | 0.0002 | 1 | compared |
| MIMIC-III | erythrocyte_sedimentation_rate | esr | 646 | 646 | 65 | 65 | 0 | 1 | compared |
| MIMIC-III | fibrinogen | fgn | 3,414 | 3,401 | 280 | 280 | 0.002 | 1 | compared |
| MIMIC-III | fio2 | fio2 | 65,349 | 65,304 | 50 | 50 | 0.0002 | 1 | compared |
| MIMIC-III | glucose | glu | 75,005 | 67,400 | 122 | 121 | 0.0097 | 0.0024 | registered definitional difference |
| MIMIC-III | hba1c | hba1c | 367 | 367 | 6 | 6 | 0 | 1 | compared |
| MIMIC-III | heart_rate | hr | 577,287 | 577,449 | 86 | 86 | 0.0001 | 1 | compared |
| MIMIC-III | hematocrit | hct | 61,238 | 61,175 | 29.6 | 29.6 | 0.0001 | 1 | compared |
| MIMIC-III | hemoglobin | hgb | 52,033 | 51,744 | 10.1 | 10.1 | 0.0006 | 1 | compared |
| MIMIC-III | lactate | lact | 17,291 | 17,245 | 1.7 | 1.7 | 0.0004 | 1 | compared |
| MIMIC-III | lymphocytes | lymph | 8,447 | 8,409 | 10.8 | 10.7 | 0.0007 | 1 | compared |
| MIMIC-III | magnesium | mg | 49,718 | 49,610 | 2 | 2 | 0.0003 | 1 | compared |
| MIMIC-III | map | map | 560,116 | 560,764 | 79.33 | 79 | 0.0002 | 1 | compared |
| MIMIC-III | mch | mch | 51,575 | 51,575 | 30.4 | 30.4 | 0 | 1 | compared |
| MIMIC-III | mchc | mchc | 51,591 | 51,591 | 33.7 | 33.7 | 0 | 1 | compared |
| MIMIC-III | mcv | mcv | 51,572 | 51,572 | 90 | 90 | 0 | 1 | compared |
| MIMIC-III | methemoglobin | methb | 306 | 306 | 0.1 | 0.1 | 0 | 1 | compared |
| MIMIC-III | mgcs | mgcs | 159,898 | 159,964 | 6 | 6 | 0.0001 | 1 | compared |
| MIMIC-III | neutrophils | neut | 8,417 | 8,409 | 80.4 | 80.4 | 0.0004 | 1 | compared |

| | | | | | | | | | |
|---|---|---|---|---|---|---|---|---|---|
| MIMIC-III | o2sat | o2sat | 567,683 | 605,820 | 98 | 98 | 0.0382 | <0.0001 | registered definitional difference |
| MIMIC-III | pco2 | pco2 | 38,984 | 38,984 | 41 | 41 | 0 | 1 | compared |
| MIMIC-III | ph | ph | 41,239 | 41,239 | 7.4 | 7.4 | 0 | 1 | compared |
| MIMIC-III | phosphate | phos | 45,905 | 45,872 | 3.3 | 3.3 | 0.0001 | 1 | compared |
| MIMIC-III | platelet | plt | 52,944 | 52,887 | 231 | 231 | 0.0002 | 1 | compared |
| MIMIC-III | po2 | po2 | 38,250 | 38,250 | 115 | 116 | 0 | 1 | compared |
| MIMIC-III | potassium | k | 64,179 | 57,747 | 4 | 4 | 0.0086 | 0.0229 | registered definitional difference |
| MIMIC-III | prothrombin_time_inr | inr_pt | 31,160 | 31,076 | 1.2 | 1.2 | 0.0002 | 1 | compared |
| MIMIC-III | pt | pt | 31,137 | 31,077 | 13.8 | 13.8 | 0.0003 | 1 | compared |
| MIMIC-III | ptt | ptt | 33,079 | 32,999 | 31.2 | 31.2 | 0.0004 | 1 | compared |
| MIMIC-III | rass | rass | 33,391 | 33,405 | 0 | 0 | 0.0001 | 1 | compared |
| MIMIC-III | rbc | rbc | 51,577 | 51,577 | 3.34 | 3.34 | 0 | 1 | compared |
| MIMIC-III | rdw | rdw | 51,504 | 51,504 | 15 | 15 | 0 | 1 | compared |
| MIMIC-III | resp_rate | resp | 606,565 | 653,910 | 19 | 18 | 0.0358 | <0.0001 | registered definitional difference |
| MIMIC-III | sbp | sbp | 557,682 | 558,417 | 123 | 123 | 0.0002 | 1 | compared |
| MIMIC-III | so2 | sao2 | 8,255 | 26,366 | 97 | 97 | 0.1302 | <0.0001 | registered definitional difference |
| MIMIC-III | sodium | na | 56,638 | 56,582 | 139 | 139 | 0.0001 | 1 | compared |
| MIMIC-III | spo2 | spo2 | 559,428 | 582,260 | 98 | 98 | 0.0379 | <0.0001 | compared |
| MIMIC-III | temperature | temp | 150,123 | 232,420 | 37.11 | 37.17 | 0.0198 | <0.0001 | registered definitional difference |
| MIMIC-III | totalco2 | tco2 | 38,985 | 38,985 | 26 | 26 | 0 | 1 | compared |
| MIMIC-III | troponin_t | tnt | 4,398 | 3,534 | 0.06 | 0.08 | 0.145 | <0.0001 | registered definitional difference |
| MIMIC-III | vgcs | vgcs | 160,199 | 160,266 | 1 | 1 | 0.0002 | 1 | compared |
| MIMIC-III | wbc | wbc | 51,988 | 51,898 | 9.9 | 9.9 | 0.0002 | 1 | compared |
| MIMIC-IV | albumin | alb | 31,283 | 31,179 | 3.1 | 3.1 | 0.0007 | 1 | compared |
| MIMIC-IV | alp | alp | 54,472 | 54,281 | 103 | 103 | 0.0005 | 1 | compared |
| MIMIC-IV | alt | alt | 55,118 | 54,645 | 26 | 26 | 0.0049 | 0.5249 | compared |
| MIMIC-IV | ast | ast | 55,687 | 55,471 | 31 | 31 | 0.0013 | 1 | compared |
| MIMIC-IV | band | bnd | 10,692 | 10,637 | 1 | 1 | 0.0009 | 1 | compared |
| MIMIC-IV | base_excess | be | 56,460 | 55,697 | 0 | 0 | 0.0004 | 1 | compared |
| MIMIC-IV | basos | basos | 39,084 | 38,929 | 0.2 | 0.2 | 0.0005 | 1 | compared |

| | | | | | | | | | |
|---|---|---|---|---|---|---|---|---|---|
| MIMIC-IV | bicarbonate | bicar | 234,339 | 233,679 | 25 | 25 | 0.0002 | 1 | compared |
| MIMIC-IV | bili | bili | 53,997 | 53,618 | 0.5 | 0.5 | 0.0032 | 0.9479 | compared |
| MIMIC-IV | bili_dir | bili_dir | 2,145 | 2,060 | 1 | 1.1 | 0.023 | 0.6328 | compared |
| MIMIC-IV | bun | bun | 235,701 | 235,022 | 18 | 18 | 0.0004 | 1 | compared |
| MIMIC-IV | c_reactive_protein | crp | 7,499 | 7,476 | 53.4 | 53.4 | 0.0009 | 1 | compared |
| MIMIC-IV | calcium_ionized | cai | 30,648 | 30,257 | 1.12 | 1.12 | 0.0004 | 1 | compared |
| MIMIC-IV | calcium_total | ca | 205,602 | 205,008 | 8.7 | 8.7 | 0.0004 | 1 | compared |
| MIMIC-IV | chloride | cl | 246,747 | 237,955 | 102 | 102 | 0.0074 | <0.0001 | compared |
| MIMIC-IV | creatine_kinase | ck | 17,233 | 17,110 | 123 | 122 | 0.0027 | 1 | compared |
| MIMIC-IV | creatine_kinase_mb | ckmb | 13,583 | 13,382 | 4 | 4 | 0.0078 | 0.8076 | compared |
| MIMIC-IV | creatinine | crea | 236,349 | 235,648 | 0.9 | 0.9 | 0.0002 | 1 | compared |
| MIMIC-IV | dbp | dbp | 916,088 | 891,202 | 62 | 62 | 0.0044 | <0.0001 | compared |
| MIMIC-IV | egcs | egcs | 257,273 | 257,273 | 4 | 4 | 0 | 1 | compared |
| MIMIC-IV | eosinophils | eos | 39,079 | 38,924 | 1 | 1 | 0.0009 | 1 | compared |
| MIMIC-IV | erythrocyte_sedimentation_rate | esr | 1,975 | 1,972 | 60 | 60 | 0.0008 | 1 | compared |
| MIMIC-IV | etco2 | etco2 | 16,830 | 16,830 | 36 | 36 | 0 | 1 | compared |
| MIMIC-IV | fibrinogen | fgn | 8,376 | 8,278 | 316 | 316 | 0.0018 | 1 | compared |
| MIMIC-IV | fio2 | fio2 | 143,367 | 143,332 | 40 | 40 | 0.0001 | 1 | compared |
| MIMIC-IV | glucose | glu | 254,698 | 251,087 | 115 | 115 | 0.0026 | 0.3417 | compared |
| MIMIC-IV | hba1c | hba1c | 3,383 | 3,383 | 5.9 | 5.9 | 0 | 1 | compared |
| MIMIC-IV | heart_rate | hr | 911,313 | 911,313 | 84 | 84 | 0 | 1 | compared |
| MIMIC-IV | hematocrit | hct | 238,812 | 238,138 | 30.2 | 30.2 | 0.0004 | 1 | compared |
| MIMIC-IV | hemoglobin | hgb | 227,459 | 226,687 | 9.9 | 9.9 | 0.0004 | 1 | compared |
| MIMIC-IV | lactate | lact | 39,178 | 38,575 | 1.6 | 1.6 | 0.0015 | 1 | compared |
| MIMIC-IV | lymphocytes | lymph | 39,239 | 38,930 | 14.1 | 14.1 | 0.0006 | 1 | compared |
| MIMIC-IV | magnesium | mg | 213,614 | 213,003 | 2 | 2 | 0.0005 | 1 | compared |
| MIMIC-IV | map | map | 915,812 | 916,927 | 79 | 79 | 0.0002 | 1 | compared |
| MIMIC-IV | mch | mch | 225,406 | 225,406 | 29.9 | 29.9 | 0 | 1 | compared |
| MIMIC-IV | mchc | mchc | 225,421 | 225,421 | 32.6 | 32.6 | 0 | 1 | compared |
| MIMIC-IV | mcv | mcv | 225,411 | 225,411 | 91 | 91 | 0 | 1 | compared |
| MIMIC-IV | methemoglobin | methb | 266 | 266 | 0 | 0 | 0 | 1 | compared |
| MIMIC-IV | mgcs | mgcs | 256,241 | 256,241 | 6 | 6 | 0 | 1 | compared |
| MIMIC-IV | neutrophils | neut | 39,084 | 38,929 | 73.55 | 73.5 | 0.0011 | 1 | compared |
| MIMIC-IV | o2sat | o2sat | 912,527 | 1,006,157 | 97 | 97 | 0.0824 | <0.0001 | registered definitional difference |
| MIMIC-IV | pco2 | pco2 | 55,721 | 55,721 | 42 | 42 | 0 | 1 | compared |
| MIMIC-IV | ph | ph | 59,117 | 59,117 | 7.39 | 7.39 | 0 | 1 | compared |
| MIMIC-IV | phosphate | phos | 205,712 | 205,115 | 3.4 | 3.4 | 0.0003 | 1 | compared |

| | | | | | | | | | |
|---|---|---|---|---|---|---|---|---|---|
| MIMIC-IV | platelet | plt | 228,529 | 227,877 | 221 | 221 | 0.0002 | 1 | compared |
| MIMIC-IV | po2 | po2 | 52,550 | 52,550 | 105 | 109 | 0 | 1 | compared |
| MIMIC-IV | potassium | k | 241,690 | 241,012 | 4.1 | 4.1 | 0.0001 | 1 | compared |
| MIMIC-IV | prothrombin_time_inr | inr_pt | 105,147 | 104,650 | 1.2 | 1.2 | 0.0003 | 1 | compared |
| MIMIC-IV | pt | pt | 105,151 | 104,661 | 13.6 | 13.6 | 0.0003 | 1 | compared |
| MIMIC-IV | ptt | ptt | 111,192 | 110,699 | 33.1 | 33.1 | 0.0005 | 1 | compared |
| MIMIC-IV | rass | rass | 176,737 | 176,737 | 0 | 0 | 0 | 1 | compared |
| MIMIC-IV | rbc | rbc | 225,414 | 225,414 | 3.35 | 3.35 | 0 | 1 | compared |
| MIMIC-IV | rdw | rdw | 225,289 | 225,289 | 14.9 | 14.9 | 0 | 1 | compared |
| MIMIC-IV | resp_rate | resp | 964,866 | 1,130,165 | 19 | 19 | 0.0363 | <0.0001 | registered definitional difference |
| MIMIC-IV | sbp | sbp | 916,950 | 891,407 | 120 | 120 | 0.0024 | 0.0106 | compared |
| MIMIC-IV | so2 | sao2 | 13,100 | 21,173 | 96 | 96 | 0.0867 | <0.0001 | registered definitional difference |
| MIMIC-IV | sodium | na | 241,638 | 240,946 | 138 | 138 | 0.0007 | 1 | compared |
| MIMIC-IV | spo2 | spo2 | 899,427 | 984,984 | 97 | 97 | 0.0842 | <0.0001 | registered definitional difference |
| MIMIC-IV | temperature | temp | 250,195 | 250,205 | 36.94 | 36.94 | 0 | 1 | compared |
| MIMIC-IV | totalco2 | tco2 | 55,720 | 55,720 | 26 | 26 | 0 | 1 | compared |
| MIMIC-IV | troponin_t | tnt | 12,176 | 10,283 | 0.06 | 0.08 | 0.1387 | <0.0001 | registered definitional difference |
| MIMIC-IV | vgcs | vgcs | 256,880 | 256,880 | 4 | 4 | 0 | 1 | compared |
| MIMIC-IV | wbc | wbc | 226,422 | 225,608 | 8.6 | 8.6 | 0.0007 | 1 | compared |

**Supplementary Table 8:** KS Concepts' differences.

| **Source** | **Concept** | **Expected shape** | **Reason** |
|---|---|---|---|
| MIMIC-III | chloride | superset | rendehR adds blood-gas chloride (50806) and charted chloride that does not copy a lab row; ricu reads labevents 50902 only |
| MIMIC-III | glucose | superset | rendehR adds charted glucose (811/1529/220621/226537) that does not copy a lab row; ricu reads labevents only |
| MIMIC-III | potassium | superset | rendehR adds charted potassium that does not copy a lab row; ricu reads labevents 50971 only |
| MIMIC-III | troponin_t | superset | rendehR adds charted troponin T (851/227429); ricu reads labevents 51003 only |

| | | | |
|---|---|---|---|
| MIMIC-IV | troponin_t | superset | rendehR adds charted troponin T (227429): charted 0.01 values of lab results reported as '<0.01' (NULL in labevents); ricu reads labevents 51003 only |
| MIMIC-III | o2sat | subset | ricu's o2sat adds SaO2 chart item 834 and SpO2 alarm limit 226253 |
| MIMIC-IV | o2sat | subset | ricu's o2sat adds SpO2 alarm limit 226253 |
| MIMIC-III | spo2 | subset | ricu's spo2 adds 226253 (SpO2 desaturation alarm limit) |
| MIMIC-IV | spo2 | subset | ricu's spo2 adds 226253 (SpO2 desaturation alarm limit) |
| MIMIC-III | resp_rate | subset | ricu's resp adds ventilator set and spontaneous rates (619/224688/224689); rendehR keeps them in resp_rate_setting / out |
| MIMIC-IV | resp_rate | subset | ricu's resp adds ventilator set and spontaneous rates (224688/224689) |
| MIMIC-III | so2 | subset | ricu's sao2 adds charted SaO2 (834/220227); rendehR so2 is the blood-gas lab value |
| MIMIC-IV | so2 | subset | ricu's sao2 adds charted SaO2 (220227); rendehR so2 is the blood-gas lab value |
| MIMIC-III | temperature | subset | ricu keeps both the Celsius and the Fahrenheit charting of the same minute; rendehR keeps one (repeated chart rows) |
| eICU-CRD | sbp | superset | rendehR adds non-invasive blood pressure (vitalaperiodic); ricu reads invasive vitalperiodic only |
| eICU-CRD | dbp | superset | rendehR adds non-invasive blood pressure (vitalaperiodic); ricu reads invasive vitalperiodic only |
| eICU-CRD | heart_rate | superset | rendehR adds nursecharting heart rate; ricu reads vitalperiodic only |
| eICU-CRD | resp_rate | superset | rendehR adds nursecharting respiratory rate; ricu reads vitalperiodic only |
| eICU-CRD | glucose | subset | ricu's glu includes bedside glucose; rendehR keeps it in glucose_fingerstick |
| eICU-CRD | calcium_ionized | time_only | eICU reports ionized calcium in mg/dL (labmeasurenamesystem); rendehR converts to mmol/L; ricu reads the mixed labmeasurenameinterface labels and keeps few rows |
| eICU-CRD | erythrocyte_sedimentation_rate | ricu_absent | ricu defines no eICU item |
| eICU-CRD | etco2 | ricu_absent | ricu defines no eICU item |
| MIMIC-III | hbco | ricu_absent | ricu reads no MIMIC-III carboxyhemoglobin row on the demo |
| MIMIC-IV | hbco | ricu_absent | ricu reads no MIMIC-IV carboxyhemoglobin row on the demo |
| eICU-CRD | insulin_exp | dur_superset | rendehR counts insulin orders (medication) and infusions; ricu's ins reads infusions only |
| eICU-CRD | epinephrine_exp | dur_superset | rendehR counts epinephrine orders and infusions; ricu's epi_dur reads infusions only |

| | | | |
|---|---|---|---|
| eICU-CRD | norepinephrine_exp | dur_superset | rendehR counts norepinephrine orders and infusions; ricu's norepi_dur reads infusions only |
| eICU-CRD | phenylephrine_exp | dur_superset | rendehR counts phenylephrine orders and infusions; ricu's phn_rate reads infusions only |
| eICU-CRD | dopamine_exp | dur_superset | rendehR counts dopamine orders and infusions; ricu's dopa_dur reads infusions only |
| eICU-CRD | vasopressin_exp | dur_superset | rendehR counts vasopressin orders and infusions; ricu's adh_rate reads infusions only |
| MIMIC-IV | insulin_exp | dur_superset | rendehR counts every insulin input (subcutaneous included); ricu's ins reads intravenous insulin only |
| eICU-CRD | map | superset | rendehR adds nursecharting MAP (invasive and non-invasive); ricu reads vitalperiodic only |
| MIMIC-IV | po2 | superset | ricu drops pO2 below its plausibility minimum of 40 mmHg (venous samples); rendehR keeps them |
| MIMIC-III | dopamine_exp | dur_subset | ricu's dopa_rate includes CareVue item 30125 which is milrinone (d_items); rendehR reads 30043/30307/221662 only |
| MIMIC-III | insulin_exp | dur_superset | rendehR counts every insulin input (subcutaneous included); ricu's ins reads intravenous insulin only |
| MIMIC-III | weight_admit | static_def | rendehR takes the first valid weight of a pooled admit/daily/echo series (documented decision in flat/weight_admit.sql); ricu reads Admit Wt items only |
| eICU-CRD | weight_admit | static_def | at hospital-stay level ricu averages admissionweight over the unit stays; rendehR takes the admission weight of the first unit stay |
| MIMIC-III | calcium_ionized | superset | rendehR adds charted ionized calcium (816/3766/225667) that does not copy a lab row; ricu reads labevents 50808 only |
| MIMIC-IV | calcium_ionized | superset | rendehR adds charted ionized calcium (225667) that does not copy a lab row; ricu reads labevents 50808 only |
| MIMIC-IV | chloride | superset | rendehR adds blood-gas chloride (50806) and charted chloride that does not copy a lab row; ricu reads labevents 50902 only |
| MIMIC-IV | creatine_kinase_mb | superset | rendehR adds charted CK-MB (227445); ricu reads labevents only |
| MIMIC-III | creatine_kinase_mb | superset | rendehR adds charted CK-MB; ricu reads labevents only |
| MIMIC-III | height | static_def | rendehR pools all real height sources (chartevent inch/cm items including 226730 |
| MIMIC-IV | weight_admit | static_def | rendehR takes the first valid weight of a pooled admit/daily/echo series at hospital level (documented decision in flat/weight_admit.sql); ricu reads its admit-weight items only |
| MIMIC-IV | phenylephrine_exp | dur_superset | rendehR counts the premixed phenylephrine formulations (229630, 229632) as well as 221749; ricu reads 221749 only (all 287 extra exposed admissions carry no 221749 rows) |

#### Supplementary code

```
# ==============================================================================
# First-day mortality in spinal cord injury, MIMIC-III + MIMIC-IV pooled.
#
# Logistic regression on first-24 h blood biomarkers, 70/30 train/test split
# on the pooled cohort. Outputs a held-out ROC curve and a log-odds-ratio
# forest plot.
# ==============================================================================

library(rendehR)
library(pROC)
library(dplyr)
library(ggplot2)

set.seed(20261103)

SCHEMA        <- "rendehr"                    # merge_sources() output, SCI cohort
OUTCOME       <- "hospital_mortality"
SOURCES       <- c("mimic_iii", "mimic_iv")
TEST_FRAC     <- 0.30
FIRST_DAY_MIN <- 1440                         # first 24 h after hospital admission
PANEL         <- c("lactate", "creatinine", "bun", "wbc", "hemoglobin", "platelet",
                   "sodium", "potassium", "bicarbonate", "glucose", "bili")

# Database credentials (or use ehr_use_config() with a project config).
CREDS <- list(postgres = list(host = "localhost", port = 5432,
                              user = "postgres", password = "…"))
ehr_connect(db_name = "rendehr", creds = CREDS, config_path = NULL)

# ---- 1. Cohort: SCI emergency admissions, MIMIC-III and MIMIC-IV ------------
```

```r
flat <- load_merged_flat(SCHEMA) |>
  filter(admission_type == "emergency", source_data %in% SOURCES)

# ---- 2. First-day labs -----------------------------------------------------
# Harmonized concepts, units already converted. Readings outside each concept's
# plausible range are dropped, then one first-day mean per admission is
# computed in the database; one row per admission comes back to R.
labs <- load_merged_ts_wide(SCHEMA, spec = PANEL, bin_min = FIRST_DAY_MIN,
                            window = c(0, 0), clean = "range")

# Alternative: pull the first day long, clean and aggregate in R.
# Useful to inspect individual readings; moves every reading into R.
# labs <- load_merged_ts(SCHEMA, concepts = PANEL, cols = "minimal",
#                        window_min = c(0, FIRST_DAY_MIN)) |>
#   mutate(value = suppressWarnings(as.numeric(value))) |>
#   clean_by_range(action = "na") |>
#   filter(!is.na(value)) |>
#   group_by(ehr_id, source_data, src_admission_id, concept_name) |>
#   summarise(value = mean(value), .groups = "drop") |>
#   tidyr::pivot_wider(names_from = concept_name, values_from = value)

dat <- inner_join(flat, labs, by = c("ehr_id", "source_data", "src_admission_id")) |>
  mutate(
    age = as.numeric(age),
    # flat stores the outcome as text; glm needs numeric 0/1
    !!OUTCOME := as.integer(as.character(.data[[OUTCOME]]) %in% c("1", "TRUE", "true", "T", "Y", "yes")),
    sex = factor(sex)
  ) |>
  mutate(across(any_of(PANEL), as.numeric)) |>
  select(source_data, all_of(OUTCOME), age, sex, any_of(PANEL)) |>
  na.omit()

stopifnot(is.numeric(dat[[OUTCOME]]), all(dat[[OUTCOME]] %in% 0:1))
table(dat$source_data, dat[[OUTCOME]])   # both classes present in each source

# ---- 3. 70/30 split on the pooled cohort, stratified by outcome -------------
test_id <- unlist(lapply(split(seq_len(nrow(dat)), dat[[OUTCOME]]),
```

```
                        function(ix) sample(ix, round(TEST_FRAC * length(ix)))))
train <- dat[-test_id, ]
test  <- dat[ test_id, ]

# Standardize with TRAINING means/SDs -> odds ratios are per 1 SD.
num <- intersect(c("age", PANEL), names(dat))
mu  <- sapply(train[num], mean); s <- sapply(train[num], sd)
std <- function(d) { d[num] <- sweep(sweep(d[num], 2, mu), 2, s, "/"); d }
train <- std(train); test <- std(test)

# ---- 4. Logistic regression ------------------------------------------------
form <- reformulate(c("age", "sex", setdiff(num, "age")), OUTCOME)
fit  <- glm(form, data = train, family = binomial())

# ---- 5. ROC on the held-out 30% --------------------------------------------
roc <- pROC::roc(test[[OUTCOME]], predict(fit, test, type = "response"))
plot(roc, print.auc = TRUE)
pROC::ci.auc(roc)

# ---- 6. Log odds ratios (forest plot) --------------------------------------
broom::tidy(fit, conf.int = TRUE) |>
  filter(term != "(Intercept)") |>
  ggplot(aes(estimate, reorder(term, estimate), xmin = conf.low, xmax = conf.high)) +
  geom_vline(xintercept = 0, linetype = 2) + geom_pointrange() +
  labs(x = "log odds ratio (per 1 SD)", y = NULL)
```
